# Circulating extracellular vesicle lipidomics identifies distinct signatures of amyotrophic lateral sclerosis and spinal muscular atrophy

**DOI:** 10.64898/2026.09.23.26363372

**Authors:** Valeria Ricotti, Antigone Fogel, Michiel Vandenbosch, Federica Cerri, Pierpaolo Ala, Alexander Capstick, Rachele Rossi, Francesca Gerardi, Ruben Jacobs, Eugenio Mercuri, Valeria Sansone, Pamela J. Shaw, Payam Barnaghi, Thomas Voit

## Abstract

Amyotrophic lateral sclerosis (ALS) is a heterogeneous neurodegenerative disorder lacking established biomarkers of underlying disease biology. Here, we identify lipidomic signatures of circulating extracellular vesicles (EVs) that distinguish ALS from neurologically healthy controls and related neuromuscular diseases. Using mass spectrometry-based profiling and multivariate modelling, we separate ALS (n=77) from healthy controls (n=89) with an area under the receiver operating characteristic curve (AUROC) of 0.90 (95% CI 0.85–0.94), rising to 0.92 (0.89-0.96) when low-confidence predictions are withheld, and retaining performance in an independent cohort (n=119; AUROC 0.78, 0.70-0.86). In Tofersen-treated SOD1-ALS patients, biomarker trajectories change in parallel with clinical measures while individuals with spinal muscular atrophy (n=50) exhibit a lipid fingerprint distinct from that observed in ALS, supporting disease specificity within a shared systemic framework of abnormal systemic lipogenesis. These findings establish EV-associated lipid signatures as a systemic, blood-based biomarker platform for ALS diagnosis, stratification, and treatment monitoring, with structurally distinct lipid species indicating widespread membrane remodelling.

## INTRODUCTION

Amyotrophic lateral sclerosis (ALS) is a neurodegenerative disorder characterised by the progressive degeneration of upper and lower motor neurons, leading to muscle weakness, functional decline and respiratory failure^1^. Although classically defined by motor neuron loss, ALS is characterised by substantial clinical and biological heterogeneity, and median survival from symptom onset remains approximately three to five years^1^. This heterogeneity has hindered therapeutic development and the identification of biomarkers that accurately reflect underlying disease biology.

Genetic and molecular studies have identified over 30 ALS-associated genes, including *SOD1, C9orf72, TARDBP* and *FUS*, implicating pathogenic mechanisms such as impaired protein homeostasis, RNA metabolism dysfunction, mitochondrial abnormalities, axonal transport defects and neuroinflammation^2,3^. While these discoveries have advanced mechanistic understanding, they have not translated into broadly effective disease-modifying therapies, highlighting a disconnect between molecular insight and clinical impact.

Increasingly, ALS is recognised not solely as a disorder of motor neurons, but as a systemic disease involving cellular dysfunction across multiple tissues and cell types^4–6^. Metabolic abnormalities, altered energy homeostasis, mitochondrial dysfunction and impaired redox balance have been reported in neurons, glial cells, skeletal muscle, immune cells and peripheral tissues in ALS, as well as in Alzheimer’s disease (AD) and Parkinson’s disease (PD)^4,7–9^. These findings indicate that systemic metabolic dysregulation may represent a core feature of neurodegeneration rather than a secondary consequence of neuronal loss.

Metabolic impairment is increasingly viewed as a unifying pathogenic axis across neurodegenerative diseases. Disruption of cellular bioenergetics, metal homeostasis, oxidative stress responses and macromolecular metabolism have been implicated in disease initiation and progression, often preceding overt neurodegeneration^7–10^. Disturbances in metal metabolism and metal-dependent enzymatic processes have been linked to mitochondrial dysfunction, altered redox signalling and impaired protein folding, processes central to neurodegenerative pathology^10,11^. This systemic perspective raises the possibility that early disease biology can be detected in peripheral biofluids, before irreversible neuronal injury occurs.

Therapeutic progress in ALS has historically been modest. Riluzole and edaravone confer limited survival or functional benefit, and numerous investigational agents have failed in late-stage clinical trials^12^. However, the therapeutic landscape has shifted recently, with increased emphasis on biologically targeted, mechanism-driven approaches. Advances highlight growing momentum in antisense oligonucleotide therapies, RNA-targeting strategies, and metabolic and immune-modulating interventions, alongside improved trial design and patient stratification^13^.

In this context, tofersen, which targets SOD1 mRNA, has received regulatory approval primarily on the basis of biomarker modulation rather than unequivocal clinical efficacy^14^. While these developments represent important progress towards precision medicine in ALS, they underscore the need for biomarkers that capture core disease biology, enable early and biologically informed patient stratification, and provide sensitive pharmacodynamic readouts beyond downstream measures of neuroaxonal injury.

A major barrier to therapeutic development is the absence of biomarkers that enable early diagnosis, stratify patients, monitor disease progression, and serve as sensitive pharmacodynamic readouts. Clinical endpoints such as the ALS Functional Rating Scale-Revised (ALSFRS-R) and survival remain essential for characterising clinical evolution but are relatively insensitive to short-term biological change and early or pre-symptomatic disease stages^15^.

The ALS biomarker landscape has expanded substantially in recent years. Among fluid-based biomarkers, neurofilament light chain (NfL), measured in cerebrospinal fluid and blood, has emerged as the most robust and widely validated marker of neuroaxonal injury, with prognostic value and correlation with disease progression^16,17^. However, neurofilaments primarily reflect downstream neuronal damage, lack disease specificity, and exhibit limited dynamic range once disease is established^17^. As such, they provide limited insight into upstream pathogenic processes, systemic metabolic dysfunction or therapeutic mechanisms of action.

Beyond neurofilaments, biomarker candidates include inflammatory mediators, metabolic indicators, RNA and microRNA species, extracellular vesicles and multi-omics signatures^17–19^. Recent plasma proteomics studies have identified candidate biomarker panels predictive of ALS, supporting high-dimensional peripheral profiling approaches^20^. Whole-blood gene expression signatures derived from high-throughput transcriptomic profiling can accurately predict ALS case status and survival, using machine learning to integrate multi-feature data, underscoring the potential of complex peripheral biomarker patterns to reflect disease biology beyond single-analyte measures^21,22^. While these approaches have generated insights, most studies have focused on isolated analytes rather than integrated biomarker signatures capable of capturing the multifactorial and systemic nature of neurodegenerative disease biology.

Importantly, the growing recognition of ALS and related neurodegenerative disorders as systemic diseases suggests that circulating biomarkers reflecting global cellular metabolism and homeostasis may provide earlier and more informative readouts than neuron-specific markers. Blood-based biomarkers are attractive, offering minimally invasive access to disease-relevant biology and enabling longitudinal monitoring at scale^23^.

Here, we report the identification and characterisation of exosomal lipid biomarker signatures in ALS and spinal muscular atrophy (SMA). As a genetically defined motor neuron disease with clinical features overlapping ALS but a distinct molecular aetiology and disease course, SMA provides a relevant comparator for assessing the disease specificity of such systemic biomarker signatures^24^.

The Vesalic platform was developed to capture integrated biomarker patterns in peripheral biofluids that may reflect systemic biological alterations. Under this framework, the signatures are explored as indicators of altered cellular metabolism across tissues, rather than direct measures of neuronal loss.

To test this hypothesis, we analysed well-characterised patient cohorts using standardised, serum-based exosomal lipid extraction quantitatively assessed by mass spectrometry, combined with multivariate analysis and machine learning across unbiased lipidomic biomarker panels. We demonstrate that Vesalic biomarker signatures robustly distinguish individuals with ALS from neurologically healthy controls and from SMA, supporting disease specificity despite shared motor neuron involvement. The high accuracy of these signatures further supports their diagnostic potential. Moreover, the structure of these signatures is consistent with sensitivity to early and potentially pre-symptomatic metabolic dysfunction, providing a quantitative basis for risk stratification that extends beyond binary labels. Longitudinal observations provide preliminary evidence of biomarker modulation in response to approved antisense ALS therapy, suggesting that these fingerprints may function as dynamic indicators of therapeutic response. Collectively, these findings position our biomarkers as a novel, systemic, blood-based platform for disease stratification, early detection and treatment monitoring across neurodegenerative disorders affecting motor neurons.

## METHODS

### Data Source & Study Cohorts

Human blood samples were obtained from 216 participants (Supplementary Table S1) recruited across three specialised clinical centres and a commercial biospecimen provider (BioIVT), comprising 77 patients with ALS, 89 neurologically healthy controls (HC), and 50 patients with spinal muscular atrophy (SMA), including 25 individuals with SMA type 2 and 25 with SMA type 3. The independent validation cohort comprised 66 individuals with ALS and 53 neurologically healthy controls (Supplementary Table S2).

All participants provided written informed consent. Ethical approval was obtained from the relevant institutional review boards or independent ethics committees at each participating site (Supplementary Table S3), including Fondazione Policlinico Universitario Agostino Gemelli IRCCS (protocol RF-2019-12370334), Centro Clinico NeMO (protocol 3929_S_N), the Northern Ireland Motor Neurone Disease Biobank (protocol 21/NI/0010), and the Sheffield Institute for Translational Neuroscience (SITraN) ALS/MND Biosample Repository (protocol 12/YH/0330).

Demographic and clinical data, including age, sex, disease duration, treatment status, and clinical scores, were collected where available. In ALS patients, clinical assessment included the ALS Functional Rating Scale-Revised (ALSFRS-R)^15^, Medical Research Council (MRC) muscle strength grading^25^, and forced vital capacity (FVC). In SMA patients, functional status (for example, sitting ability) and motor performance were evaluated using the Hammersmith Functional Motor Scale-Expanded (HFMSE)^26^ and the Revised Upper Limb Module (RULM)^27^.

Participants were recruited through neurology clinics, patient registries, clinical trials, and biobanking services. ALS diagnosis was established according to the El Escorial criteria^28^, while controls had no history of neurological or neuromuscular disease. Genetic testing for ALS-associated variants was performed according to standard clinical diagnostic workflows, including targeted gene panels or next-generation sequencing approaches to detect pathogenic variants in genes such as SOD1, C9orf72, TARDBP and FUS, alongside repeat-primed PCR for hexanucleotide repeat expansions in C9orf72^29^. SMA diagnosis was confirmed by genetic testing demonstrating homozygous deletion or mutation of the SMN1 gene, using established methods such as multiplex ligation-dependent probe amplification (MLPA) or quantitative PCR to assess SMN1 copy number and distinguish it from SMN2^30^.

Both treatment-naïve and treated patients were included; treatment status was recorded and accounted for in downstream analyses. Exclusion criteria comprised the presence of other neurological or neurodegenerative disorders, active systemic infection or inflammatory disease at the time of sampling, or malignancy.

All procedures were conducted in accordance with the Declaration of Helsinki and are reported in line with the STROBE Statement guidelines^31^.

### Sample Preparation and Exosome Isolation

Blood samples were collected in anticoagulant-free tubes and permitted to clot at room temperature, then centrifuged at 2,000 × g for 10 minutes to remove cellular debris. Serum was aliquoted and stored at −80°C until exosome isolation. Exosomes were isolated from serum using the Total Exosome Isolation Reagent (Thermo Fisher Scientific, 4478360) according to the manufacturer’s protocol, with minor modifications. Briefly, 150 µl of serum was centrifuged at 2,000 × g for 30 minutes at 4°C to remove large particles. The supernatant was transferred to a new tube and incubated with 30 µl of the isolation reagent on ice for 30 minutes. After centrifugation at 10,000 × g for 10 minutes at room temperature, the pellet was resuspended in 50 µl of phosphate-buffered saline (PBS) and stored at −80°C for lipidomic analysis.

### Lipid Extraction and LC–MS Analysis

Lipid extraction from exosome pellets was performed using a modified Bligh-Dyer method^32^. A 25 µl aliquot of internal standard (SPLASH® Lipidomix®; Avanti Polar Lipids) was added to 25 µl of exosome suspension, followed by vortex mixing. The samples were then diluted with 200 µl of ultrapure water and centrifuged at 13,000 × g for 10 minutes at 4°C. The organic lipid phase was isolated and dried under vacuum. Lipids were reconstituted in 50 µl of methanol:chloroform (1:2, v/v) and stored at −80°C until analysis. The extraction process was validated using quality control (QC) samples prepared from pooled serum, ensuring consistency across batches and minimising variability in lipid recovery.

The LC–MS lipidomics was conducted using a Thermo Scientific Ultimate 3000 UHPLC system coupled to a Q Exactive HF Orbitrap mass spectrometer (Thermo Scientific) according to a generic lipidomics protocol^33^. Lipids were separated on a Hypersil GOLD C18 column (100 × 2.1 mm, 1.9 µm) with a gradient elution protocol: mobile phase A (acetonitrile:water, 60:40, v/v, 10 mM ammonium formate) and mobile phase B (isopropanol:acetonitrile, 90:10, v/v, 10 mM ammonium formate). The gradient was optimised to separate lipid classes. Full MS scans (m/z 200–1,450, resolution 60,000) and data-dependent MS/MS scans were acquired in positive ion mode. Lipid species were annotated using Lipostar2 software (v2.1.7) and quantified by normalising peak areas to internal standards. The lipidomic analysis was performed in triplicate for QC samples to ensure reproducibility, and the detection limit for each lipid class was determined based on the signal-to-noise ratio.

## Bioinformatics and Statistical Analysis

### Data preprocessing

Samples were analysed in five batches, with only the first run retained for samples analysed more than once. ALS and control samples were distributed evenly in three batches with a small number of control samples (n=19) run in 2 boxes that did not contain any ALS samples (Supplementary Table S4). The initial untargeted lipidomic analysis identified more than 100 lipids. Only lipids present in all batches were retained, and any lipid feature containing a missing value was removed (n=3), yielding a complete matrix of 35 lipids across 166 samples with no missing or zero entries and thus no imputation of undetected values performed. The same 35 lipids were quantified in the external validation cohort (n=119). Intensities were log2-transformed and each feature was z-score standardised once across the full dataset for univariate analysis and within a scikit-learn Pipeline for predictive modelling.

### Statistical analysis

To assess statistical differences in lipid intensities between disease groups, the non-parametric Mann-Whitney U and Kruskal-Wallis tests were used due to the non-normal distribution of lipid intensity measures^34,35^. Pairwise differences in lipid intensities within the same individuals before and after treatment were evaluated using the Wilcoxon signed-rank test^36^. For all statistical analyses across lipid features, multiple testing was controlled using the Benjamini–Hochberg false discovery rate (FDR) correction to limit false-positive findings while maintaining statistical power across the large number of comparisons^37^.

### Predictive modelling

Predictive performance was evaluated using two supervised learning models selected for their utility in clinical prediction tasks and complementary modelling characteristics: logistic regression (LR) and eXtreme Gradient Boosting (XGBoost)^38^. LR assumes that each lipid contributes additively to the log-odds of disease and provides directly interpretable coefficients. XGBoost, a gradient-boosted tree ensemble, relaxes that assumption and can represent non-linear feature-to-outcome relationships and interactions between lipids. Comparing the two tests whether disease-associated information in this panel is additive or requires higher-order structure.

Model training and evaluation were performed using a nested cross-validation approach with five outer folds and five inner folds implemented with stratified sampling and sample shuffling *(StratifiedKFold*) to preserve class balance across folds and minimise sampling bias (Figure 1b). The inner loop was used for hyperparameter tuning, while the outer loop was used for unbiased estimation of predictive performance. Within each inner loop, models were implemented as a pipeline consisting of z-score standardisation, feature selection (*SelectFromModel* for XGBoost; *SelectKBest* for logistic regression), and classification. The number of selected features was fixed at 25 across models to reduce overfitting while preserving performance based on preliminary experiments across a range of feature set sizes. Hyperparameters were optimised for average precision using a randomised search strategy. Performance metrics, including F1 score, accuracy, precision, sensitivity, specificity, area under the precision-recall curve (AUPRC), and area under the receiver operating characteristic curve (AUROC), were calculated for each fold. Results were summarised as the mean across folds with corresponding 95% confidence intervals.

**Figure 1.**
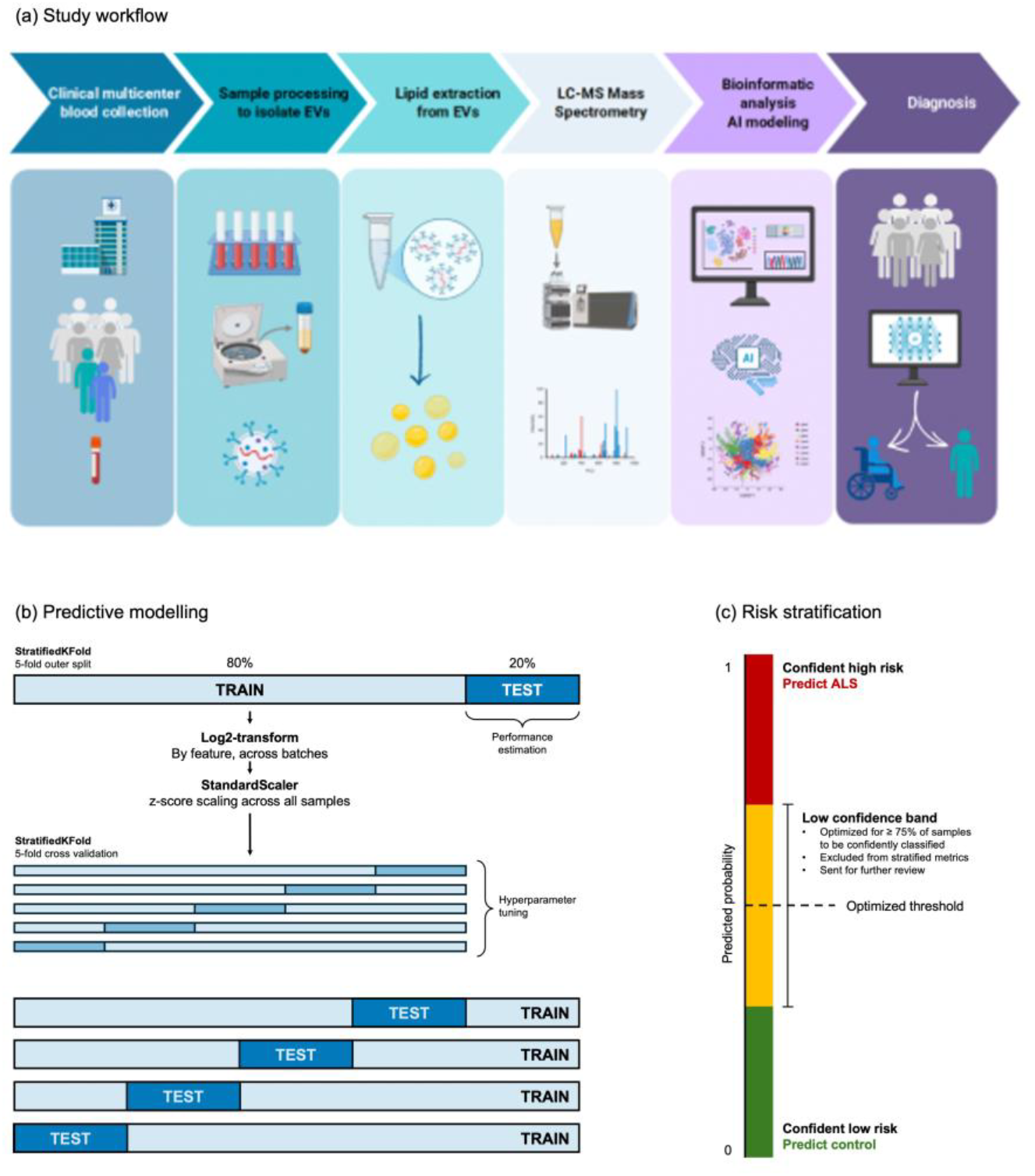
Overview of Vesalic biomarker platform. (a) Study workflow and analytical framework. Blood samples were collected across multiple clinical sites from individuals with ALS, disease comparators, and healthy controls. Extracellular vesicles (EVs) were isolated from serum and subjected to high-resolution lipidomic profiling using liquid chromatography–mass spectrometry. Lipid abundance measures were analysed using statistical analysis and predictive modelling to identify disease-associated signatures. ML-derived continuous disease probability scores enable diagnostic discrimination and risk stratification. Created in BioRender. Rossi, R (2026) . (b) Predictive modelling approach involved nested cross validation with 5 inner folds for hyperparameter optimisation and 5 outer folds for performance estimation. (c) Predicted probabilities of ALS were stratified into confident high and low risk categories, with an intermediate low confidence band excluded from stratified predictions.

Models were trained using only lipid features from the full ALS and control participant dataset (n = 166).

### Risk stratification

To increase confidence in model predictions, a selective classification approach was implemented (Figure 1c)^39^. An uncertainty band was defined surrounding the decision threshold such that samples with predicted probabilities close to the threshold, where model confidence is reduced, were classified as uncertain rather than being assigned a definitive class label. The uncertainty band width was optimised to maximise classification performance for confidently classified samples while ensuring at least 75% of samples received a definitive classification. Samples were thus stratified into three risk categories: low (Green), high (Red), and uncertain (Amber). Both the decision threshold and the band width were determined within the training partition of each outer fold, using out-of-fold predicted probabilities from an internal 5-fold cross-validation, and evaluation metrics were calculated after applying the thresholds unchanged to the held-out test data.

### Model explainability

To quantify the contribution of individual features to predicted risk, models were retrained on the full dataset, and Shapley Additive exPlanations (SHAP) values were computed^40^. Global feature importance was assessed using summary plots that capture both the magnitude and direction of feature influence across the dataset.

### Evaluating treatment response

To evaluate the sensitivity of lipid biomarkers to longitudinal treatment responses, the best-performing model was retrained on the full dataset, excluding all longitudinal samples from treated individuals. The trained model was then applied to longitudinal samples to generate predicted probabilities of ALS at each time point. Risk stratification thresholds derived from the training data were applied to these predictions, and predicted probabilities were visualised over time for each individual.

## RESULTS

### Circulating EV lipidomics identify a reproducible ALS-associated signature across sporadic and monogenic disease

To investigate whether circulating extracellular vesicle (EV)-associated lipid profiles capture disease-relevant biology in ALS, we developed an integrated analytical framework combining standardised blood collection across multiple clinical sites, extracellular vesicle isolation from serum, and high-resolution lipidomic profiling using liquid chromatography-mass spectrometry, followed by multivariate analysis and training a predictive model (Figure 1a). This approach was applied to well-characterised cohorts including both sporadic and monogenic ALS, alongside healthy controls and disease comparators.

We then assessed whether the resulting lipid signatures could (i) distinguish ALS from controls, (ii) define a continuous disease probability landscape, (iii) reflect dynamic biological changes in response to therapy, and (iv) differentiate ALS from related neuromuscular disorders.

The primary ALS dataset comprised 166 individuals, including 77 patients with ALS and 89 neurologically healthy controls (Supplementary Table S1). The ALS cohort included both sporadic (n=44) and monogenic (n=29) cases (n=4 unknown), enabling the analysis to capture a broader representation of ALS biology rather than a genetically restricted subtype. This is particularly relevant for biomarker development, as clinically useful platforms must detect convergent disease-associated biology across heterogeneous aetiologies, rather than perform only within narrowly defined molecular subgroups.

We first examined whether circulating exosomal lipid profiles contain a disease-associated signal that distinguishes ALS from healthy controls.

Initial univariate analysis identified 25 lipids that differed significantly between ALS and healthy control samples after Benjamini-Hochberg correction for multiple comparisons (Figure 2 and Supplementary Table S5). Across these features, the direction and magnitude of the group differences were consistent with a coordinated disease-associated shift rather than isolated outlier behaviour. Although individual feature effect sizes varied, the overall pattern was notable for its coherence across multiple lipids, supporting the view that ALS is associated with a reproducible change in circulating exosomal lipid homeostasis detectable in blood-derived EV fractions. The distributions shown in Figure 2 and quantified in Supplementary Table S5 indicate that many of these lipids exhibit partial group overlap when considered individually but nevertheless display clear shifts in median abundance and distributional shape between ALS and controls. This pattern strongly suggests that diagnostic performance is unlikely to arise from any single feature in isolation and instead depends on the aggregate information embedded across the signature fingerprint as a whole.

**Figure 2.**
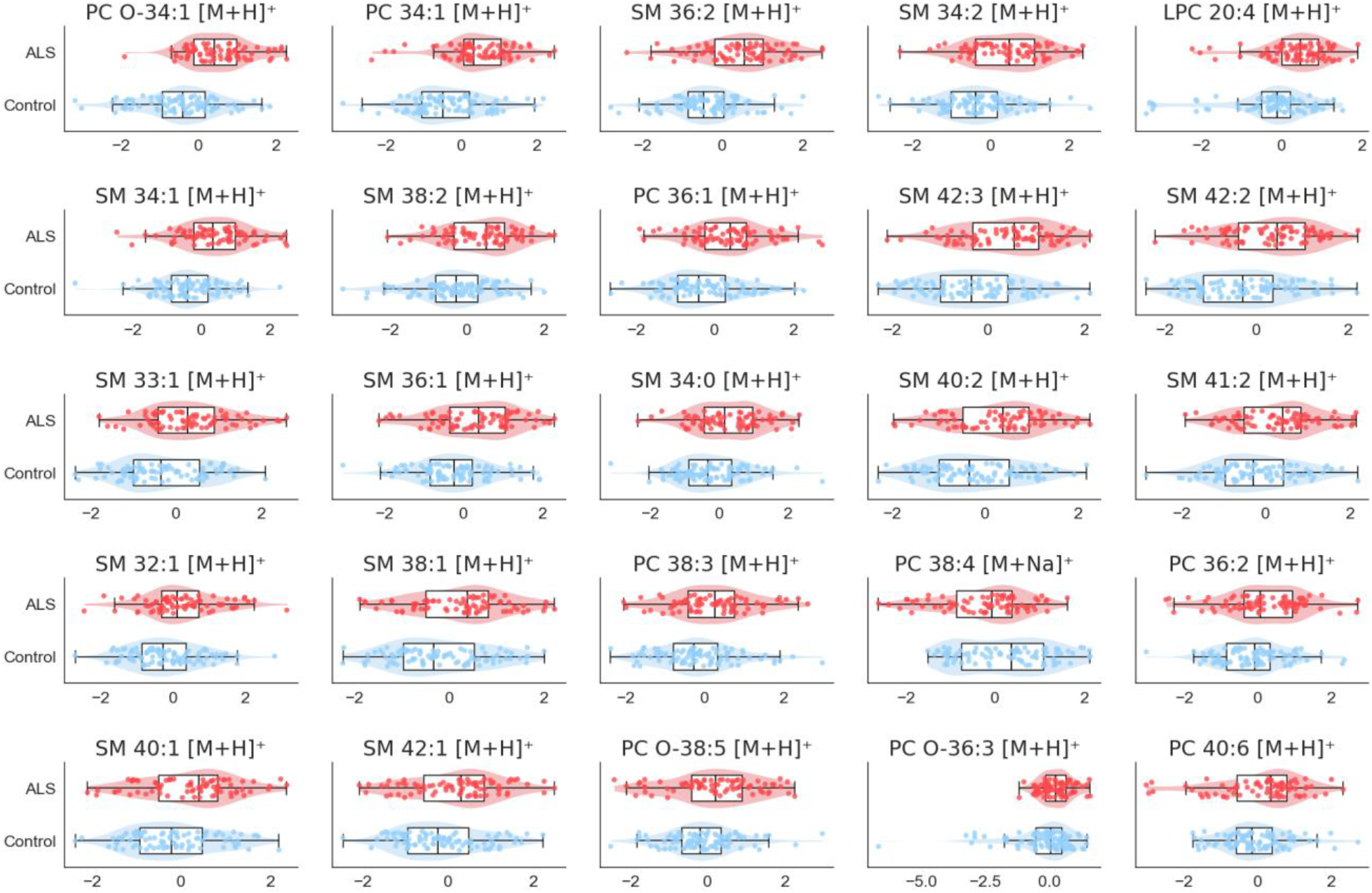
Differential lipidomic feature distributions in ALS. Distribution of the 25 lipids that differ significantly between ALS patients (n=77) and healthy controls (n=89), as measured by a Mann-Whitney U test with Benjamini-Hochberg FDR correction for multiple comparisons. Individual lipid features display moderate but consistent shifts in abundance between groups, with substantial overlap at the single-analyte level. These patterns indicate that disease-associated differences are distributed across multiple lipid species rather than driven by a single dominant biomarker. Raw lipid values were log2-transformed and z-score standardised prior to plotting.

ALS appears to introduce a structured perturbation superimposed on a background of inter-individual variation. Although overall separation between groups is limited, consistent differences are observed across multiple features, indicating that disease-associated information is not reflected in a dominant global shift but rather in coordinated variation across a subset of variables.

This feature of the data is particularly relevant in the context of ALS heterogeneity. Both sporadic and monogenic ALS samples contributed to a shared broad region of discriminative signal despite differing upstream causes, and only one lipid differed significantly between these groups, suggesting the presence of shared biological features across the ALS spectrum (see Supplementary Figure S1 and Supplementary Table S6).

### Circulating lipidomic signatures define a continuous ALS risk landscape with high predictive accuracy

A logistic regression model trained on circulating lipid features accurately discriminated between ALS and neurologically healthy controls across the cohort (n = 166), outperforming the comparative XGBoost model (Figure 3c, Supplementary Table S7, and Supplementary Figure S2). The model achieved an area under the receiver operating characteristic curve (AUROC) of 0.90 (95% CI: 0.85-0.94), with a sensitivity of 0.77 (95% CI: 0.67-0.86) and a specificity of 0.84 (95% CI: 0.75-0.93) (Fig. 3a-c and Supplementary Table S7).

**Figure 3.**
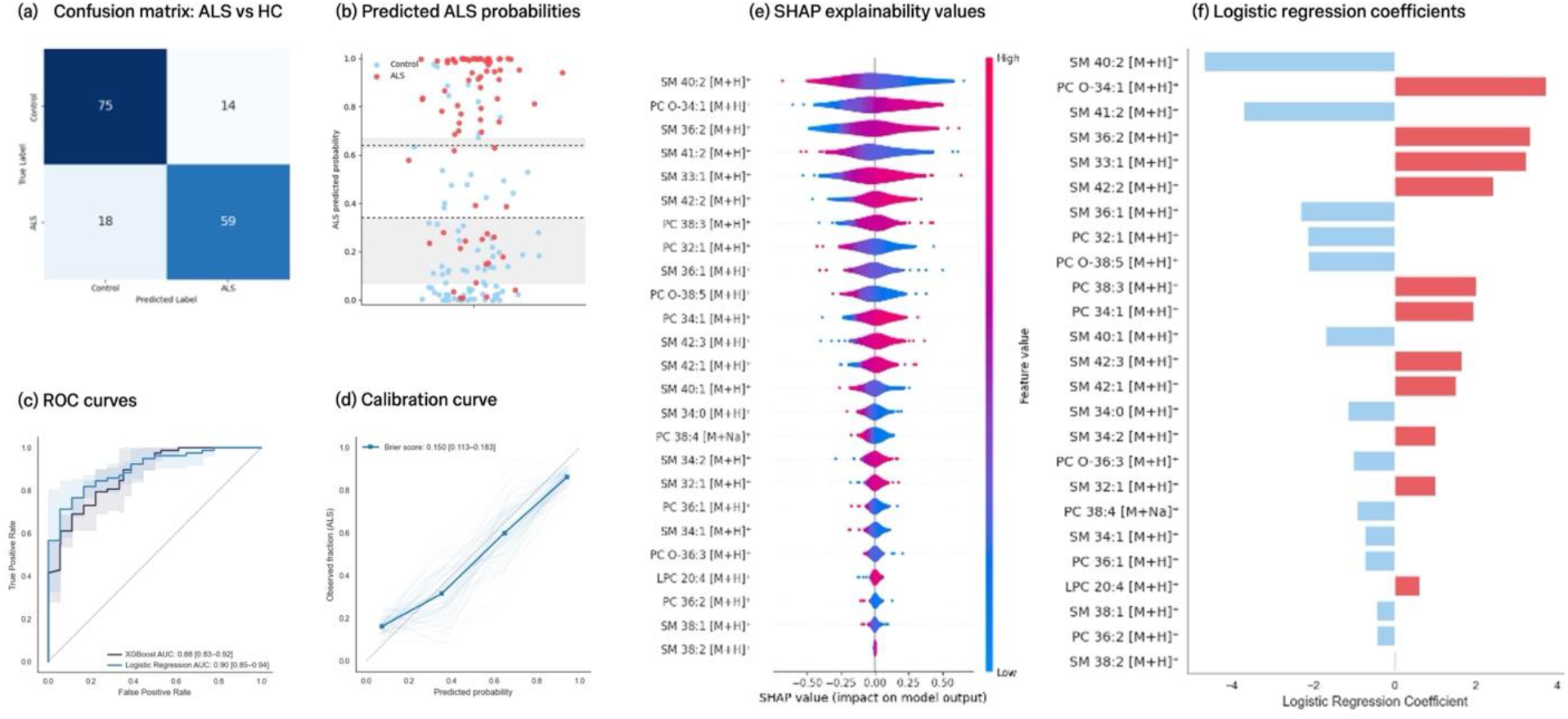
Multivariate modelling enables accurate ALS prediction and risk stratification. Performance and interpretability of the predictive model for classification of ALS versus controls using circulating lipid features across the full cohort. (a) Confusion matrix summarising classification performance. Counts of true positives, true negatives, false positives, and false negatives are shown for the model. (b) Risk stratification analysis showing classification of individuals into high-confidence ALS, high-confidence control, and intermediate probability groups. Each point represents a sample, its colour corresponds to the true disease state of that sample, and the y-axis corresponds to the biomarker-derived predicted probability of ALS. Dashed lines represent median upper and lower thresholds, and grey shaded regions represent the interquartile range across 5 outer folds. (c) Receiver operating characteristic (ROC) curves demonstrating high discriminative performance of the LR and XGBoost models. (d) Calibration curve for ALS classification showing observed ALS fraction against predicted probabilities, binned across all predictions. The solid line represents the pooled calibration curve, while faint lines show 100 bootstrap resamplings. (e) SHAP (SHapley Additive exPlanations) summary plot illustrating the contribution of individual lipid features to model predictions. Features are ranked by importance, with colour indicating relative feature value and x-axis representing the direction of effect on predicted risk. (f) Logistic regression coefficients demonstrate the magnitude and direction of each feature’s contribution to the model’s predictions.

Receiver operating characteristic (ROC) curves demonstrated consistent separation between ALS and control samples across cross-validation folds, with limited variability in performance, supporting the internal stability of the classifier (Figure 3c). Precision-recall analyses were similarly robust, indicating reliable performance even under conditions of potential class imbalance. At the level of individual predictions, the model achieved a balanced trade-off between sensitivity and specificity, correctly classifying the majority of ALS and control samples (0.81 (95% CI: 0.73-0.88), as reflected in the confusion matrix (Fig. 3a).

Beyond binary classification, the model generated continuous predicted probability scores, positioning individuals along an ALS risk continuum. Predicted probabilities showed clear separation and reasonable calibration between ALS and control populations, with most ALS samples assigned high predicted probabilities and controls clustering at low values (Fig. 3b and d). Samples falling near the decision threshold corresponded to lower-confidence predictions, defining a transition region between classes. Stratification based on predicted risk enabled classification into high-confidence, high-risk, and low-risk groups, as well as an intermediate, uncertain category. Following risk stratification, AUROC, sensitivity, and specificity of the model increased to 0.92 (95% CI: 0.89-0.96), 0.83 (95% CI: 0.71-0.95), and 0.85 (95% CI: 0.69-1.00), respectively, with 79% of individuals in outer fold test sets classified with high confidence.

From a clinical perspective, this predictive framework provides a more nuanced representation of disease state than binary classification, capturing graded variation across the disease continuum. This approach supports identification of individuals in intermediate or early disease states and has potential utility for diagnostic workflows.

To investigate the biological basis of model predictions, feature contributions were analysed using SHapley Additive exPlanations (SHAP) values (Figure 3e) alongside logistic regression coefficients (Figure 3f). ALS classification was not driven by a single dominant lipid species, but by the combined effect of multiple features, each contributing directionally to the predicted probability of disease. The lipid features spanned multiple classes, including phosphatidylcholines (PCs), sphingomyelins (SMs), and lysophosphatidylcholines (LPCs), with individual species contributing in opposing directions to disease probability. This pattern suggests a redistribution of lipid species rather than uniform class-level changes.

While univariate analyses suggest a general increase in lipid abundance in ALS, the multivariate model reveals a structured pattern: features contribute with opposing signs, such that predicted probability reflects the balance across multiple lipids rather than abundance of any single lipid. Because the model is additive on the log-odds scale, these effects are conditional on the other features and not statistical interactions between them, with several coefficients opposing their univariate association. This pattern is consistent with coordinated alterations in lipid metabolism, whereby relative changes across lipid species reflect shifts in metabolic pathways. The biomarker captures a reorganisation of cellular lipid homeostasis, distributed across multiple organs and encoded in the circulating EV lipid profile, rather than isolated changes in individual molecules.

The distribution and ranking of SHAP values were consistent with a high-dimensional molecular architecture in which combined feature effects contribute to diagnostic performance (Figure 3e). The consistency of feature contributions across samples supports a shared disease-associated molecular state. Because the ALS cohort includes both sporadic and monogenic cases, this indicates that the model captures convergent downstream biology rather than genotype-specific effects.

The multi-feature structure of the biomarker confers robustness relative to single-analyte approaches. Biomarkers based on individual molecules often reflect downstream neuronal injury and may lack sensitivity to early or systemic disease processes. In contrast, a coordinated lipidomic signature may capture broader aspects of cellular homeostasis, reflecting exosome-derived signals from multiple tissues. Circulating extracellular vesicle lipid signatures therefore provide a continuous ALS risk landscape, characterised by high predictive accuracy, stable performance across cohorts, and a distributed molecular architecture consistent with systemic disease biology.

### Longitudinal Tofersen data suggest that biomarker trajectories capture individual treatment responsiveness

We next assessed whether lipid biomarkers respond dynamically to Tofersen, an antisense oligonucleotide that downregulates SOD1 expression^14^. Longitudinal samples from seven Tofersen-treated ALS patients (median age [range]: 57 [46-64], M/F: 5/2) provided an opportunity to assess whether model-derived ALS probabilities change over time in parallel with clinical evolution.

Post-treatment biomarker trajectories and model-derived probabilities changed within individuals over the sampling period, and the direction and magnitude of change differed between patients (Fig. 4a-d). Under the pre-specified model, three of seven patients moved from a high risk profile at baseline to an intermediate (n=2) or low-risk (n=1) profile following treatment suggesting a potential measurable therapy-associated shift at the biomarker level. Four participants (49, 50, 53, and 54; Figure 4 a-d) showed particularly clear concordance between clinical and biomarker-derived changes, in both directions. In participants 49 and 53, improvements in ALSFRS and FVC were accompanied by a reduction in model-derived ALS probability. In participants 50 and 54, reduction in ALSFRS and FVC were accompanied by increases in model-derived ALS probability following treatment initiation; N60 received the lowest clinical scores and highest model-derived probability at 6 months post-treatment. These observations are descriptive and were not formally tested given the small number of participants with available longitudinal treatment data.

**Figure 4.**
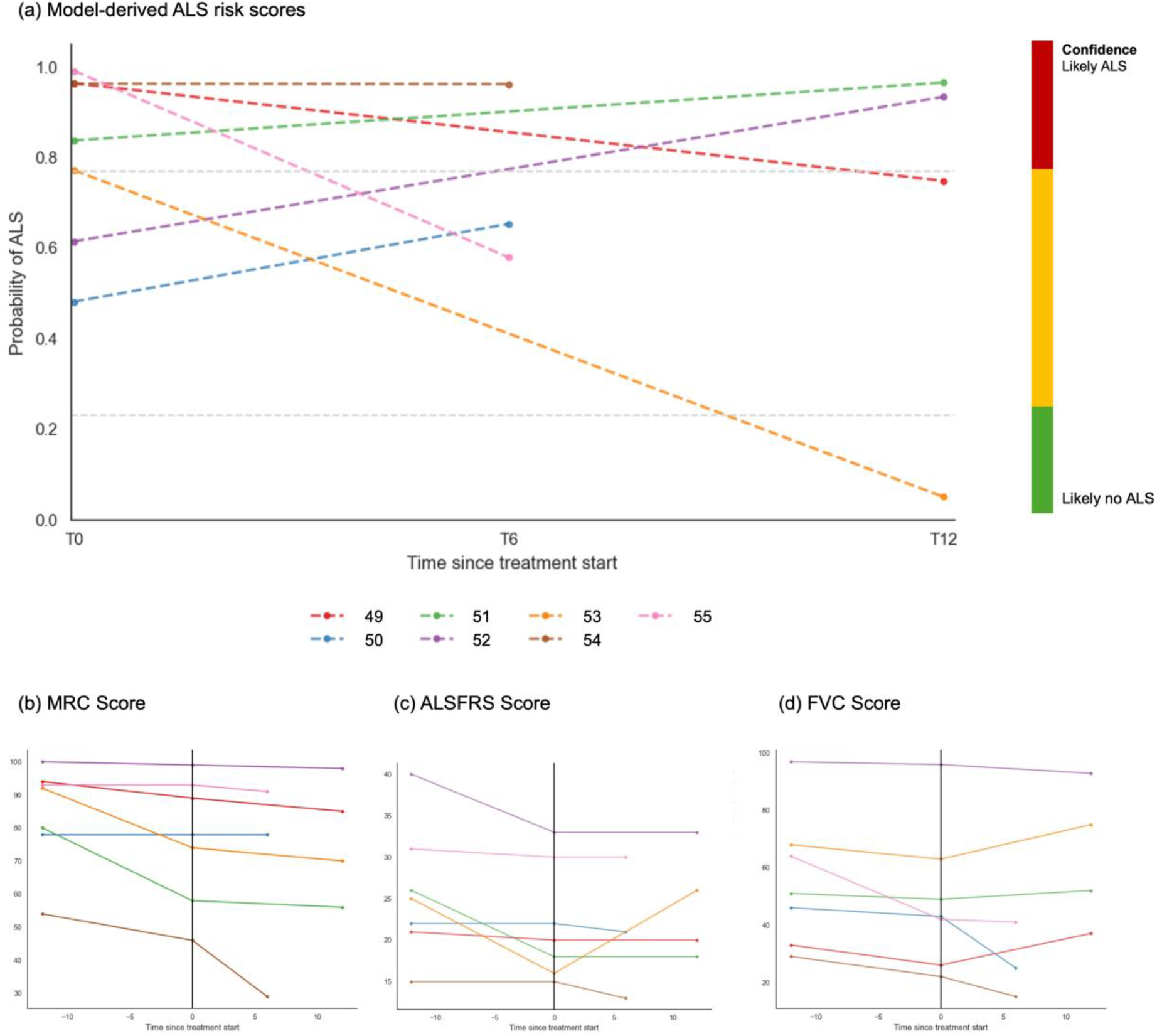
Longitudinal biomarker dynamics and clinical trajectories in Tofersen-treated patients. Longitudinal analysis of seven ALS patients treated with Tofersen, showing model-derived biomarker risk trajectories alongside clinical measures over time. In panels b-d, treatment initiation is indicated by the vertical line. (a) Model-derived ALS probability (biomarker prediction risk) over time for individual patients, demonstrating temporal modulation of disease risk and inter-individual variability following treatment. (b) Medical Research Council (MRC) muscle strength scores over time. (c) Amyotrophic Lateral Sclerosis Functional Rating Scale–Revised (ALSFRS-R) scores over time. (d) Forced vital capacity (FVC) over time. On all three clinical scales, a lower score suggests greater impairment. Across panels, trajectories show heterogeneous patterns between patients. Alignment between biomarker and clinical measures supports the dynamic and biologically relevant nature of the circulating lipidomic signature.

Longitudinally, biomarker trajectories followed a continuous, directional pattern from baseline (T0) through 6 and 12 months, rather than exhibiting oscillatory behaviour (Figure 4a). This pattern is consistent with a coordinated, system-level response to treatment, suggestive of progressive reorganisation of underlying metabolic processes rather than stochastic fluctuations.

At the individual level, biomarker dynamics mirrored clinical heterogeneity. For example, the patient with the highest baseline muscle strength (Medical Research Council (MRC) score^25^) showed a gradual increase in predicted disease probability over time (T0-T6-T12), in parallel with a slow but progressive decline in clinical function, as reflected by ALS Functional Rating Scale-Revised (ALSFRS-R) scores^15^ and forced vital capacity (FVC) (Figure 4b-d). This observation suggests a sensitivity of the biomarker to subtle disease progression, even in cases with relatively preserved baseline function.

Consistent with the multivariate nature of the model, biomarker trajectories reflect the combined contribution of multiple lipid species rather than changes in individual analytes. The presence of distinct, patient-specific trajectories indicates that the biomarker captures inter-individual variability in disease progression and treatment response, rather than a uniform cohort-level effect. Supplementary longitudinal trajectories illustrate this variability across patients, highlighting convergent and divergent response patterns within the treated cohort (Supplementary Figures S3 and S4).

Taken together, these findings indicate that the exosomal lipidomic signature is dynamic and reflects longitudinal changes in disease-associated biology, including inter-individual variability in therapeutic response. Given the small cohort, absence of a comparator group and lack of formal statistical analysis of longitudinal trajectories, these findings should be considered descriptive and hypothesis-generating.

### SMA provides a motor neuron disease-specific control and demonstrates that the ALS signature is not a generic motor neuron pathology signal

To assess whether the observed lipidomic signature reflects disease-specific biology rather than a generic consequence of motor neuron degeneration, spinal muscular atrophy (SMA) was included as a neuromuscular disease control (Supplementary Table S1). This represents a stringent comparison, as SMA shares motor system involvement with ALS but differs fundamentally in its genetic origin and underlying pathobiology, being caused by loss-of-function of the SMN1 gene^41^.

Across individual lipid features, SMA samples exhibited a consistent and distinct distributional shift compared to both ALS and healthy controls (Figure 5a and Supplementary Figure S5). While ALS was characterised by an overall increase in lipid abundance relative to healthy controls, SMA lipid profiles were characterised by a relative reduction in lipid abundance. Analysis of SMA subtypes further reinforced this pattern. SMA type 2 and type 3 samples showed consistent directional shifts relative to controls, with a graded magnitude corresponding to clinical severity, such that SMA type 2 exhibited greater deviation than SMA type 3 (Figure 5b and Supplementary Figure S6). The coherence of these shifts across lipid features indicates that the observed alterations reflect a structured, disease-associated metabolic programme rather than stochastic variation.

**Figure 5.**
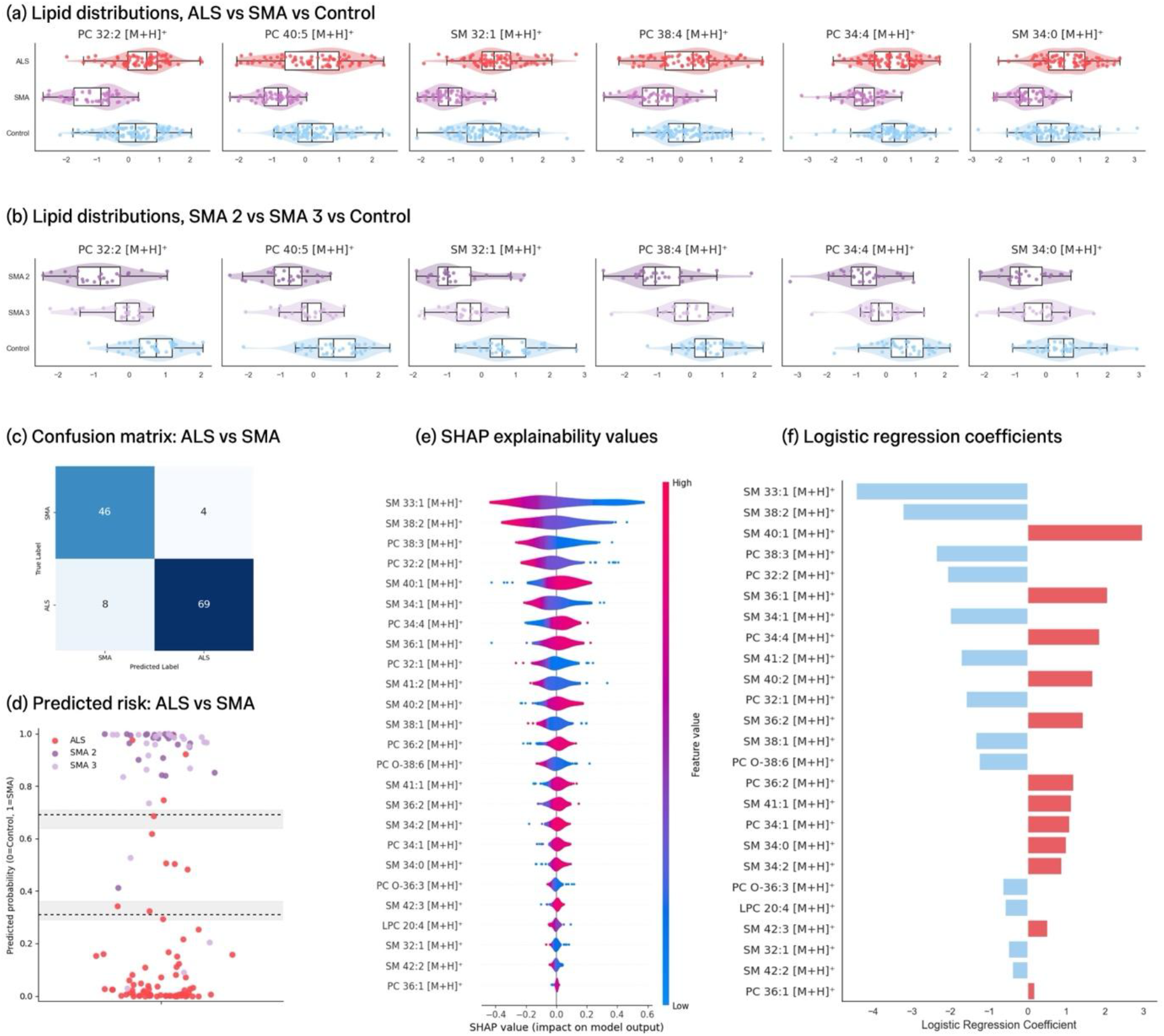
Lipidomic signatures across ALS, SMA, and control cohorts. (a) Comparison of lipidomic profiles across ALS (n=77), spinal muscular atrophy (SMA) (n=50), and healthy control (n=38) cohorts, demonstrating disease-specific molecular signatures and subtype-dependent patterns. (b) Distribution of selected lipid features across SMA2 (n=25), SMA3 (n=25), and control groups, illustrating coordinated and directionally aligned changes across lipid species, with magnitude of alteration reflecting disease severity. (c) Confusion matrix summarising classification performance for the model trained to classify SMA vs ALS. (d) Model-derived predicted probabilities of ALS vs SMA, where a probability of 0 indicates a high likelihood of ALS, and 1 indicates a high likelihood of SMA. Each prediction is coloured based on the sample’s true disease, with SMA2 and SMA3 coloured distinctly. SHAP (e) and logistic regression coefficient (f) values for the model predicting ALS vs SMA. Positive SHAP and coefficient values push predictions toward SMA, while negative SHAP and coefficient values push toward a prediction of ALS.

Applying the same machine learning pipeline, ALS and SMA samples were effectively discriminated (AUROC 0.96 (95% CI: 0.92-1.00), Sensitivity 0.92 (95% CI: 0.86-0.98), and Specificity 0.90 (95% CI: 0.83-0.97)), with low misclassification rates as illustrated by the confusion matrix (Figure 5c). Clear separation between ALS and SMA samples was further evident in predicted probability distributions, where the two groups occupied largely non-overlapping regions across both optimised threshold and risk-stratified frameworks (Figure 5d).

Importantly, this discrimination was maintained despite the majority of SMA patients (48/50) receiving SMN-enhancing therapies (nusinersen or risdiplam), indicating that treatment with nusinersen or risdiplam is not capable of fully reversing the disease-specific metabolic alteration. If the observed signal reflected a non-specific consequence of motor neuron injury, substantial overlap between ALS and SMA would be expected. Instead, the consistent separation supports the presence of disease-specific systemic molecular states.

To investigate the biological basis of discrimination between ALS and SMA, feature contributions were examined using SHAP values (Figure 5e) alongside logistic regression coefficients (Figure 5f). Classification was not driven by a single dominant lipid species, but by the combined effect of multiple features, each contributing directionally to the predicted outcome. SHAP and coefficient analyses revealed a structured pattern of feature importance, with lipid species exerting bidirectional contributions. Positive SHAP values and coefficients were associated with increased likelihood of SMA classification, while negative values favoured ALS, indicating that separation arises from the relative weighting of shared lipid features rather than the presence of entirely distinct molecules.

This multivariate structure is consistent with coordinated alterations in lipid metabolism, where relative shifts across lipid classes reflect pathway-level differences between diseases. Consistent feature contributions across samples support stable, disease-associated molecular states, while the distributed signal suggests robustness beyond single-analyte biomarkers.

Because most ALS samples were run in earlier batches than SMA samples (Supplementary Table S4), disease group is partially confounded with batch and the observed discrimination may be inflated. The pipeline was therefore rerun restricted to the ALS (n=20) and SMA (n=40) samples analysed in batch 4. Minimal changes were observed in performance metrics with AUROC and Sensitivity falling to 0.93 (95% CI: 0.84-1.00) and 0.88 (95% CI: 0.68-1.00) and Specificity rising to 0.95 (95% CI: 0.81-1.00).

Taken together, these findings indicate that ALS and SMA occupy distinct yet partially overlapping positions within a shared metabolic landscape. SMA subtypes form a graded continuum, with lipidomic perturbation scaling with disease severity, while ALS shows a reproducible signature across sporadic and monogenic cases. Despite overlapping motor phenotypes, the distinct lipidomic profiles of ALS and SMA support disease-specific organisation of cellular metabolism rather than a generic consequence of tissue damage.

### External validation cohort

To evaluate the reproducibility and generalisability of the identified EV-associated lipid signature, we applied the trained model to an independent external validation cohort comprising 119 samples from 66 individuals with ALS and 53 neurologically healthy controls collected at separate sites and processed in a different batch than any samples in the training cohort (Supplementary Table S2). The external cohort was processed using parameters fit on the training set; it did not influence model training or training set preprocessing. When applied to the external cohort, the pre-trained model achieved an AUROC of 0.78 (95% CI: 0.70-0.86) with a sensitivity of 0.65 (95% CI: 0.53-0.76), and specificity of 0.81 (95% CI: 0.71-0.91). Risk stratification retained 76% of predictions and increased performance to an AUROC of 0.81 (95% CI: 0.71-0.89), a sensitivity of 0.67 (95% CI: 0.53-0.79), and a specificity of 0.85 (95% CI: 0.74-0.95). Consistent with the findings in the primary study cohort, no significant differences were observed between the lipid fingerprints of monogenic (n=10) and sporadic (n=56) ALS in the external validation cohort.

Sensitivity and specificity confidence intervals overlapped between internal cross-validation and external validation. Performance was lower in the external cohort, as expected when a fixed model and decision threshold are applied to samples from different sites and analytical batches. Transfer to the external cohort without recalibration provides a conservative estimate of generalisability. In clinical applications, recalibration of the decision threshold to the intended-use population will be required, rather than retraining.

### Overall conclusion

The analyses show that EV-associated lipidomic biomarkers capture a characteristic molecular fingerprint with high diagnostic accuracy in ALS (Figure 2). This signature is not defined by a single analyte or a dominant global shift detectable through unsupervised approaches, but by a coordinated, multi-feature pattern resolved through multivariate modelling and expressed as continuous disease risk.

Across the cohort, lipid-only models achieved an AUROC of 0.90 (95% CI: 0.85–0.94) (Figure 3b and Supplementary Table S7). Performance remained balanced across sensitivity, specificity and precision, with the confusion matrix demonstrating correct classification of ALS and control samples (Figure 3a). Predicted risk distributions showed clear separation between groups, with a limited transition region corresponding to lower-confidence predictions (Figure 3b).

The signal was consistent across sporadic and monogenic ALS, and feature contribution analyses demonstrated that classification was driven by multiple lipid species rather than a single dominant feature (Figure 3e,f).

Longitudinal analysis of Tofersen-treated patients revealed dynamic modulation of model-derived disease probabilities, with patient-specific trajectories aligned with changes in muscle strength, functional status and respiratory function (Figure 4).

SMA subtypes exhibited consistent, directionally coherent lipidomic shifts relative to controls, while remaining distinct from ALS across distributional, probabilistic and model-based analyses (Figure 5 and Supplementary Figures S5 and S6).

Reproducibility in an independent external validation cohort supports the robustness and generalisability of the EV-associated lipid signature across patient populations and analytical batches.

Together, these findings support a model in which EV-associated lipidomic signatures reflect systemic, cell-derived signals linking peripheral alterations to disease biology. The contributions of distinct lipid species, including PCs, SMs and LPCs, are consistent with coordinated membrane remodelling rather than uniform class-level alterations.

## DISCUSSION

Our biomarker platform supports a model in which circulating extracellular vesicle (EV)-associated lipid signatures reflect a systemic and biologically structured state in ALS that extends beyond downstream markers of neuroaxonal injury. Current fluid biomarkers, most notably neurofilament light chain (NfL), have demonstrated robust diagnostic and prognostic utility in ALS, with strong correlations to disease progression and survival^16,17^. However, NfL primarily reflects neuroaxonal damage once neurodegeneration is established and therefore represents a downstream readout of disease activity rather than a direct measure of upstream pathogenic processes^16,17,42^. While highly sensitive to disease presence, NfL lacks disease specificity and provides limited insight into the biological mechanisms driving ALS or distinguishing it from related neurodegenerative and neuromuscular disorders^16,42,43^.

In contrast, the EV-associated lipidomic signatures identified here demonstrate high diagnostic accuracy and disease specificity, distinguishing ALS from healthy controls and from related conditions such as SMA types 2 and 3. Notably, the biomarker does not differ between sporadic and monogenic ALS, indicating convergence onto a shared molecular state downstream of mono- or polygenic deficits. The observation that SMA subtypes exhibit consistent yet distinct directional lipidomic shifts further supports disease-specific systemic molecular states, rather than a shared downstream signal of motor neuron degeneration. This indicates that the biomarker captures aspects of disease biology beyond neuronal injury and reflects broader alterations in cellular and metabolic homeostasis across tissues. To our knowledge, this is the first study to demonstrate that a circulating EV-associated lipidomic signature can simultaneously discriminate ALS from healthy individuals and a neuromuscular disease comparator, while showing longitudinal modulation in treated patients. Together, these findings extend EV lipidomics beyond cross-sectional biomarker discovery towards a multidimensional framework integrating disease classification, biological stratification and treatment monitoring.

The coordinated, multi-feature architecture of our biomarkers further supports this interpretation. Unlike single-analyte biomarkers, which represent isolated biological processes, the lipidomic signature reflects a distributed molecular state across multiple tissues. This is consistent with evidence that ALS is not solely a disorder of motor neurons but a multisystem disease characterised by metabolic dysregulation, mitochondrial dysfunction, and altered cellular homeostasis^4–6^. The ability of the biomarker to capture such a distributed signal suggests that it may provide a more integrated view of disease biology than markers restricted to neuronal injury. Notably, individual phosphatidylcholine (PC) and sphingomyelin (SM) species contributed in opposite directions to ALS classification, indicating that lipid class membership alone does not determine functional behaviour; instead, acyl chain length, saturation and molecular architecture influence membrane packing, vesicle dynamics and metabolic turnover, consistent with a redistribution of lipid species rather than uniform class-level changes^44^.

A key mechanistic consideration is the role of extracellular vesicles as carriers of biological information. EVs are membrane-bound particles released by virtually all cell types and mediate intercellular communication through the transfer of proteins, lipids, and nucleic acids^18^. Their potential as circulating biomarkers in ALS has gained support, although methodological heterogeneity and limited independent validation remain challenges^45,46^. Their lipid composition reflects their endosomal membrane origin and the metabolic and stress states of their cells of origin. Major lipid classes identified in this study (including SMs, PCs, and LPCs) play distinct structural and functional roles within EV membranes. SMs contribute to membrane rigidity and stability, while PCs and LPCs promote membrane flexibility and curvature^46^. Functionally, SMs regulate EV biogenesis and cargo selection, whereas LPCs facilitate vesicle interactions and act as signalling molecules, particularly in inflammatory pathways^49^. LPCs have also been implicated in transport processes across the blood-brain barrier (BBB), supporting their relevance to neurodegenerative disease biology^44^. One could speculate that altered EV lipid composition in ALS and in SMA may influence EV membrane properties such as cargo packaging, signalling, and transport.

Because EVs circulate systemically, they provide a mechanism for sampling tissue-specific pathology through peripheral biofluids. EV lipids play central roles in membrane structure, energy metabolism, redox balance, and signalling pathways^7,8^. Dysregulation of lipid metabolism, including altered cholesterol transport, has been reported in ALS^47^, supporting a potential link between lipid homeostasis and disease pathogenesis. The coordinated changes across lipid species are consistent with a reorganisation of lipid homeostasis rather than isolated perturbations, suggesting that EV lipid profiles act as integrated reporters of systemic metabolic state. The discriminatory performance achieved by combining multiple lipid species, despite opposing individual contributions, further suggests that the diagnostic information resides in the architecture of the lipidomic pattern rather than any single lipid abnormality. This represents a conceptual distinction from single-analyte biomarker approaches and may be particularly suited to a biologically heterogeneous disease such as ALS.

The detectability of these signatures in circulation indicates that disease-associated alterations extend beyond the central nervous system into peripheral compartments. Lipidomic changes have also been observed in cerebrospinal fluid in ALS^48^, supporting a systemic component to disease biology. This aligns with a growing body of evidence indicating that ALS involves widespread metabolic dysfunction across tissues^4–6^. EV-associated lipid signatures may represent a convergence point at which diverse upstream cellular processes involving protein or RNA metabolism are reflected in a measurable circulating lipidomic phenotype.

The longitudinal analyses in Tofersen-treated patients support the biological relevance of this signal. Variability in biomarker trajectories across individuals mirrors the heterogeneous clinical responses observed with Tofersen and other antisense oligonucleotide therapies ^14,20^. Such variability is expected given differences in baseline disease burden, progression rate, and timing of therapeutic intervention. While some individuals exhibit stabilisation or improvement, others continue to decline despite therapy, reflecting inter-individual variability. The ability of the lipidomic signature to capture dynamic, treatment-associated biological changes supports its potential as a pharmacodynamic marker, enabling sensitive assessment of early biological response in therapeutic development^17^. Longitudinal shifts in model-derived disease probabilities during treatment, aligned with clinical trajectories, provide preliminary evidence that the signature captures dynamic changes in disease state beyond diagnostic classification.

Alignment of model-derived disease probabilities with clinical trajectories supports sensitivity to individual treatment responses. This pharmacodynamic potential is particularly relevant in ALS, where ALSFRS-R and FVC may not capture early or subtle biological responses to therapy^14,21,22^.

The ability of the lipidomic signature to respond to treatment targeting the root cause suggests utility in identifying responders, non-responders, and intermediate phenotypes, thereby supporting precision medicine approaches. By reflecting systemic metabolic dysfunction rather than downstream neuronal loss, EV-associated lipid signatures may emerge before overt neurodegeneration, supporting their potential for pre-symptomatic or early-stage disease detection. Recent longitudinal plasma proteomic studies demonstrating molecular changes before phenoconversion in genetically at-risk individuals provide independent evidence that circulating multivariate signatures can precede clinically manifest ALS^49^. This is particularly relevant in genetically defined ALS, where at-risk individuals could be monitored for the emergence of disease-associated molecular signatures.

More broadly, a circulating, multivariate biomarker reflecting systemic disease state has implications beyond ALS. The ability to define continuous disease probability, rather than binary classification, enables individuals to be positioned along a molecular spectrum of disease. Such an approach may facilitate earlier intervention, improved patient stratification, and more sensitive detection of therapeutic effects in clinical trials.

Several limitations should be considered. The cohort size, while sufficient to demonstrate robust classification performance, remains modest relative to large-scale biomarker studies, and external validation in additional independent cohorts will be important. While the data support a systemic and metabolic interpretation of the signatures, the precise cellular and tissue origins of EV lipid alterations remain to be fully defined. Future studies integrating proteomic, transcriptomic, and functional analyses will be important to further elucidate the biological pathways underlying these observations and to determine how EV lipid signatures relate to molecular alterations across tissues and biofluids^50^.

Further investigation in presymptomatic individuals, particularly genetically defined at-risk populations, will be required to determine whether the biomarker can detect disease-associated changes prior to clinical onset. In parallel, larger longitudinal studies will be required to assess whether the EV lipid signature reflects biological responses to treatment and can serve as a pharmacodynamic or disease-monitoring biomarker.

In summary, we define a circulating EV-associated lipidomic signature that robustly discriminates ALS from healthy controls, distinguishes ALS from related neuromuscular disorders, and captures treatment-associated changes in disease biology over time. In contrast to established markers of neuroaxonal injury, and complementary to emerging proteomic and transcriptomic approaches, EV lipidomics interrogate a distinct layer of disease biology linked to membrane composition, cellular metabolism and vesicle dynamics. Collectively, our findings identify EV lipidomics as a distinct biomarker modality in ALS, integrating diagnostic discrimination, disease-specific molecular stratification and preliminary sensitivity to longitudinal therapeutic change. These findings support its potential as a platform for diagnosis, patient stratification and therapeutic monitoring alongside emerging mechanism-targeted therapies.

## Data Availability

The data used in this study are available from the corresponding author upon reasonable request.

## Code Availability

The code used to generate results presented in this study is available from the corresponding author upon reasonable request.

## Author contributions

**V.R**. secured funding, conceived and conceptualised the project, contributed to study design, data analysis and interpretation, and provided clinical and translational interpretation of the findings. She wrote the first draft of the manuscript, critically reviewed and revised the manuscript and is a co-inventor on patents arising from the work; **A.F**. contributed to the bioinformatic and computational analyses, analytical methodology and data interpretation, contributed to drafting the manuscript, and critically reviewed and revised the manuscript. She is a co-inventor on one of the patents arising from the work; **M.V**. led the mass spectrometry analyses, contributed to the interpretation of the lipidomic data and study findings, contributed to drafting the manuscript, and critically reviewed and revised the manuscript; **F.C**. contributed patient samples and associated clinical information, contributed to the clinical interpretation of the data and study findings, and critically reviewed and revised the manuscript; **P.A**. led biobank activities, including the sourcing and coordination of patient samples, and contributed to study implementation and sample management, and critically reviewed and revised the manuscript; **A.C**. contributed to the bioinformatic and computational analyses, analytical methodology and data interpretation, and critically reviewed and revised the manuscript; **R.R**. contributed to data interpretation and critically reviewed the manuscript; **F.G**. contributed patient samples and associated clinical information, contributed to the clinical interpretation of the data and study findings, and critically reviewed and revised the manuscript; **R.J**. contributed to the mass spectrometry analyses and critically reviewed and revised the manuscript; **E.M**. contributed patient samples and associated clinical information, contributed to the clinical interpretation of the data and study findings, and critically reviewed and revised the manuscript; **V.S**. contributed patient samples and associated clinical information, contributed to the clinical interpretation of the data and study findings, and critically reviewed and revised the manuscript; **P.J.S**. contributed patient samples and associated clinical information, contributed to the clinical interpretation of the data and study findings, and critically reviewed and revised the manuscript; **P.B**. contributed to the conceptualisation of the project and analytical strategy, with particular responsibility for bioinformatic and computational methodology, data analysis and interpretation, and critically reviewed and revised the manuscript; **T.V**. secured funding, conceived and conceptualised the project, contributed to study design, data analysis and clinical and translational interpretation of the findings, critically reviewed and revised subsequent versions. He is a co-inventor on patents arising from the work.

*All authors reviewed and approved the final manuscript*.

## Acknowledgements

We thank Gianpaolo Cicala (Fondazione Policlinico Universitario Agostino Gemelli IRCCS), Alex Daniel and Mays Baidoun (Sheffield Institute for Translational Neuroscience, University of Sheffield), Tim Hendriks (Maastricht University), Stephanie Duguez and William Duddy (Ulster University) for their support in facilitating access to clinical samples and associated clinical data.

## REFERENCES

1. Feldman, E. L. et al. Amyotrophic lateral sclerosis. The Lancet 400, 1363–1380 (2022).

2. Taylor, J. P., Brown, R. H. & Cleveland, D. W. Decoding ALS: from genes to mechanism. Nature 539, 197–206 (2016).

3. Hardiman, O. et al. Amyotrophic lateral sclerosis. Nat. Rev. Dis. Primer 3, 17071 (2017).

4. Dupuis, L., Pradat, P.-F., Ludolph, A. C. & Loeffler, J.-P. Energy metabolism in amyotrophic lateral sclerosis. Lancet Neurol. 10, 75–82 (2011).

5. Steyn, F. J. et al. Hypermetabolism in ALS is associated with greater functional decline and shorter survival. J. Neurol. Neurosurg. Psychiatry 89, 1016–1023 (2018).

6. Goutman, S. A. et al. Metabolomics identifies shared lipid pathways in independent amyotrophic lateral sclerosis cohorts. Brain 145, 4425–4439 (2022).

7. Wang, Y., Xu, E., Musich, P. R. & Lin, F. Mitochondrial dysfunction in neurodegenerative diseases and the potential countermeasure. CNS Neurosci. Ther. 25, 816–824 (2019).

8. Muddapu, V. R., Dharshini, S. A. P., Chakravarthy, V. S. & Gromiha, M. M. Neurodegenerative Diseases - Is Metabolic Deficiency the Root Cause? Front. Neurosci. 14, 213 (2020).

9. Barnaghi, P. et al. Applying a systemic approach that extends beyond the brain to Alzheimer’s disease pathogenesis. Commun. Med. 6, 475 (2026).

10. Barnham, K. J. & Bush, A. I. Metals in Alzheimer’s and Parkinson’s Diseases. Curr. Opin. Chem. Biol. 12, 222–228 (2008).

11. Roberts, B. R., Ryan, T. M., Bush, A. I., Masters, C. L. & Duce, J. A. The role of metallobiology and amyloid-β peptides in Alzheimer’s disease. J. Neurochem. 120, 149–166 (2012).

12. Wei, Y. et al. Current therapy in amyotrophic lateral sclerosis (ALS): A review on past and future therapeutic strategies. Eur. J. Med. Chem. 272, 116496 (2024).

13. ALS Association. 36th International Symposium on ALS/MND. (2025).

14. Miller, T. et al. Phase 1–2 Trial of Antisense Oligonucleotide Tofersen for SOD1 ALS. N. Engl. J. Med. 383, 109–119 (2020).

15. Cedarbaum, J. M. et al. The ALSFRS-R: a revised ALS functional rating scale that incorporates assessments of respiratory function. J. Neurol. Sci. 169, 13–21 (1999).

16. Gaiani, A. et al. Diagnostic and Prognostic Biomarkers in Amyotrophic Lateral Sclerosis: Neurofilament Light Chain Levels in Definite Subtypes of Disease. JAMA Neurol. 74, 525–532 (2017).

17. Benatar, M. et al. ALS biomarkers for therapy development: State of the field and future directions. Muscle Nerve 53, 169–182 (2016).

18. Verber, N. S. et al. Biomarkers in Motor Neuron Disease: A State of the Art Review. Front. Neurol. 10, (2019).

19. Gagliardi, D., Bresolin, N., Comi, G. P. & Corti, S. Extracellular vesicles and amyotrophic lateral sclerosis: from misfolded protein vehicles to promising clinical biomarkers. Cell. Mol. Life Sci. 78, 561–572 (2021).

20. Chia, R. et al. A plasma proteomics-based candidate biomarker panel predictive of amyotrophic lateral sclerosis. Nat. Med. 31, 3440–3450 (2025).

21. van Rheenen, W. et al. Whole blood transcriptome analysis in amyotrophic lateral sclerosis: A biomarker study. PLoS One 13, e0198874 (2018).

22. Zhao, Y. et al. Gene expression signatures from whole blood predict amyotrophic lateral sclerosis case status and survival. Nat. Commun. 16, 9631 (2025).

23. Sturmey, E. & Malaspina, A. Blood biomarkers in ALS: challenges, applications and novel frontiers. Acta Neurol. Scand. 146, 375–388 (2022).

24. Mercuri, E., Sumner, C. J., Muntoni, F., Darras, B. T. & Finkel, R. S. Spinal muscular atrophy. Nat. Rev. Dis. Primer 8, 52 (2022).

25. Medical Research Council. Aids to the examination of the peripheral nervous system. (1976).

26. O’Hagen, J. M. et al. An expanded version of the Hammersmith Functional Motor Scale for SMA II and III patients. Neuromuscul. Disord. 17, 693–697 (2007).

27. Mazzone, E. S. et al. Revised upper limb module for spinal muscular atrophy: Development of a new module. Muscle Nerve 55, 869–874 (2017).

28. Brooks, B. R., Miller, R. G., Swash, M., Munsat, T. L., & World Federation of Neurology Research Group on Motor Neuron Diseases. El Escorial revisited: revised criteria for the diagnosis of amyotrophic lateral sclerosis. Amyotroph. Lateral Scler. Mot. Neuron Disord. Off. Publ. World Fed. Neurol. Res. Group Mot. Neuron Dis. 1, 293–299 (2000).

29. Roggenbuck, J. et al. Evidence-based consensus guidelines for ALS genetic testing and counseling. Ann. Clin. Transl. Neurol. 10, 2074–2091 (2023).

30. Prior, T. W. Carrier screening for spinal muscular atrophy. Genet. Med. 10, 840–842 (2008).

31. Vandenbroucke, J. P. et al. Strengthening the Reporting of Observational Studies in Epidemiology (STROBE): Explanation and Elaboration. PLOS Med. 4, e297 (2007).

32. Bligh, E. G. & Dyer, W. J. A rapid method of total lipid extraction and purification. Can. J. Biochem. Physiol. 37, 911–917 (1959).

33. Hendriks, T. F. E. et al. MALDI-MSI-LC-MS/MS Workflow for Single-Section Single Step Combined Proteomics and Quantitative Lipidomics. Anal. Chem. 96, 4266–4274 (2024).

34. Mann, H. B. & Whitney, D. R. On a Test of Whether one of Two Random Variables is Stochastically Larger than the Other. Ann. Math. Stat. 18, 50–60 (1947).

35. Kruskal, W. H. & Wallis, W. A. Use of Ranks in One-Criterion Variance Analysis. J. Am. Stat. Assoc. 47, 583–621 (1952).

36. Wilcoxon, F. Individual Comparisons by Ranking Methods. Biom. Bull. 1, 80 (1945).

37. Benjamini, Y. & Hochberg, Y. Controlling the False Discovery Rate: A Practical and Powerful Approach to Multiple Testing. J. R. Stat. Soc. Ser. B Methodol. 57, 289–300 (1995).

38. Chen, T. & Guestrin, C. XGBoost: A Scalable Tree Boosting System. in Proceedings of the 22nd ACM SIGKDD International Conference on Knowledge Discovery and Data Mining 785–794 (Association for Computing Machinery, New York, NY, USA, 2016). doi:10.1145/2939672.2939785.

39. El-Yaniv, R. & Wiener, Y. On the Foundations of Noise-free Selective Classification. J. Mach. Learn. Res. 11, 1605–1641 (2010).

40. Lundberg, S. M. et al. From local explanations to global understanding with explainable AI for trees. Nat. Mach. Intell. 2, 56–67 (2020).

41. Lefebvre, S. et al. Identification and characterization of a spinal muscular atrophy-determining gene. Cell 80, 155–165 (1995).

42. Khalil, M. et al. Neurofilaments as biomarkers in neurological disorders. Nat. Rev. Neurol. 14, 577–589 (2018).

43. Verde, F., Otto, M. & Silani, V. Neurofilament Light Chain as Biomarker for Amyotrophic Lateral Sclerosis and Frontotemporal Dementia. Front. Neurosci. 15, (2021).

44. Harayama, T. & Riezman, H. Understanding the diversity of membrane lipid composition. Nat. Rev. Mol. Cell Biol. 19, 281–296 (2018).

45. Brent, A., Shirmast, P. & McMillan, N. A. J. Extracellular Vesicle Lipids and Their Role in Delivery. J. Extracell. Biol. 4, e70064 (2025).

46. Ghadami, S. & Dellinger, K. The lipid composition of extracellular vesicles: applications in diagnostics and therapeutic delivery. Front. Mol. Biosci. 10, (2023).

47. Semba, R. D. Perspective: The Potential Role of Circulating Lysophosphatidylcholine in Neuroprotection against Alzheimer Disease. Adv. Nutr. 11, 760–772 (2020).

48. Sapaly, D. et al. Dysregulation of muscle cholesterol transport in amyotrophic lateral sclerosis. Brain 148, 788–802 (2025).

49. Ran, X. et al. Longitudinal plasma proteomics predict phenoconversion to clinically manifest ALS. Nat. Med. 1–13 (2026) doi:10.1038/s41591-026-04528-x.

50. Adler, G. L., Kiernan, M. C. & Tan, R. H. The FindMNDBiomarker Program: Protein Changes in Motor Neuron Disease/Amyotrophic Lateral Sclerosis Postmortem Tissue and Biofluids. Ann. Neurol. 98, 788–800 (2025).

