## Supplementary material for "Circulating extracellular vesicle lipidomics identifies distinct signatures of amyotrophic lateral sclerosis and spinal muscular atrophy": Supplementary Material medRxiv.pdf

**Table S1.** Demographic and clinical characteristics of the primary study cohort.

| Cohort | N | Sex (M / F) | Age at onset (median [range]) | Age at sample collection (median [range]) | Genetics |
|---|---|---|---|---|---|
| ALS | 77 | 46 / 29† | 60 [22–82] | 62 [29–82] ‡ | 44 sporadic<br>29 monogenic:<br>SOD1 (n = 13),<br>TDP43 (n = 5),<br>FUS (n = 4),<br>C9ORF72 (n = 7) |
| SMA type II | 25 | 16 / 9 | N/A | 25 [2–41] | SMN1 deletion / conversion |
| SMA type III | 25 | 14 / 11 | N/A | 32 [10–60] | SMN1 deletion / conversion |
| Healthy controls | 89 | 30 / 36† | N/A | 45 [24–74]‡ | N/A |

**Abbreviations:** ALS, amyotrophic lateral sclerosis; SMA, spinal muscular atrophy; SMN1, survival motor neuron 1; SOD1, superoxide dismutase 1; TDP43, TAR DNA-binding protein 43; FUS, fused in sarcoma; C9ORF72, chromosome 9 open reading frame 72; N/A, not applicable. † Sex data were available for 66/89 healthy controls and 75/77 ALS samples; 27 controls and 2 ALS samples from Ulster Biobank had no sex recorded. ‡ Age at sample collection reported for 38/89 healthy controls and 75/77 ALS patients with available data (NEMO and BIOIVT cohorts); age data were not available for Ulster Biobank samples

**Table S2.** Demographic characteristics of the external validation cohort.

| Cohort | N | Sex (M / F) | Age at onset (median [range]) | Age at sample collection (median [range]) | Genetics |
|---|---|---|---|---|---|
| ALS | 66 | 34 / 32 | 63 [29-77] | 64 [29–80] | 56 sporadic<br>10 monogenic |
| Healthy controls | 53 | 33 / 20 | N/A | 56 [26–80] | N/A |

**Abbreviations:** ALS, amyotrophic lateral sclerosis; N/A, not applicable.

**Table S3.** Collaborating centres and biospecimen sources used in this study.

| Centre / Institution | Country | Patient serum | Healthy control serum | Ethics approval |
|---|---|---|---|---|
| NEMO, Milan | Italy | ALS | HC | Ethics approval: 3929_S_N |
| Ulster Biobank | UK | ALS | HC | Ethics approval: 21/NI/0010 |
| Fondazione Policlinico Universitario Agostino Gemelli IRCCS | Italy | SMA | — | Ethics approval: RF-2019-12370334 |
| SITraN, University of Sheffield | UK | ALS | HC | Ethics approval: 12/YH/0330 |
| BIOIVT | UK / US | — | HC | Commercially supplied |

**Abbreviations:** ALS, amyotrophic lateral sclerosis; SMA, spinal muscular atrophy; HC, healthy control; IRCCS, Istituto di Ricovero e Cura a Carattere Scientifico; —, not applicable.
**Ethics approval numbers:** Patient serum and healthy control serum columns indicate sample types contributed by each centre. BIOIVT samples were obtained commercially and are not linked to a specific ethics protocol.

**Table S4.** Sample distribution across analytical batches.

| Disease | Batch 1 | Batch 2 | Batch 3 | Batch 4 | Batch 5 | Batch 6 |
|---|---|---|---|---|---|---|
| **ALS** | 30 | 27 | 0 | 20 | 0 | 0 |
| **Control** | 30 | 20 | 1 | 20 | 18* | 18* |
| **SMA** | 0 | 0 | 0 | 40 | 0 | 10 |

*Duplicate samples. Batch 5 used for ALS vs HC model, Batch 6 used for SMA vs HC analysis.

**Table S5.** Lipids that differ significantly between ALS and Control

| Feature | *U* statistic | Effect size | p-value | p-value (FDR) |
|---|---|---|---|---|
| PC O-34:1_[M+H]+ | 1466 | -0.572158 | <0.000001 | <0.000001 |
| PC 34:1_[M+H]+ | 1640 | -0.521377 | <0.000001 | <0.000001 |
| SM 36:2_[M+H]+ | 1664 | -0.514373 | <0.000001 | <0.000001 |
| SM 34:2_[M+H]+ | 1878 | -0.451919 | 0.000001 | 0.000004 |
| LPC 20:4_[M+H]+ | 1880 | -0.451335 | 0.000001 | 0.000004 |
| SM 34:1_[M+H]+ | 1929 | -0.437035 | 0.000001 | 0.000007 |
| SM 38:2_[M+H]+ | 2038 | -0.405224 | 0.000007 | 0.000032 |
| PC 36:1_[M+H]+ | 2041 | -0.404348 | 0.000007 | 0.000032 |
| SM 42:3_[M+H]+ | 2090 | -0.390048 | 0.000015 | 0.000059 |
| SM 42:2_[M+H]+ | 2242 | -0.345688 | 0.000126 | 0.000441 |
| SM 33:1_[M+H]+ | 2289 | -0.331971 | 0.000232 | 0.000737 |
| SM 36:1_[M+H]+ | 2320 | -0.322924 | 0.000342 | 0.000997 |
| SM 34:0_[M+H]+ | 2340 | -0.317087 | 0.000437 | 0.001177 |
| SM 40:2_[M+H]+ | 2410 | -0.296658 | 0.001002 | 0.002505 |
| SM 41:2_[M+H]+ | 2430 | -0.290822 | 0.001259 | 0.002938 |
| SM 32:1_[M+H]+ | 2528 | -0.262221 | 0.003640 | 0.007961 |
| SM 38:1_[M+H]+ | 2587 | -0.245002 | 0.006592 | 0.013572 |
| PC 38:3_[M+H]+ | 2601 | -0.240916 | 0.007553 | 0.014686 |
| PC 38:4_[M+Na]+ | 4220 | 0.231577 | 0.010234 | 0.018852 |
| PC 36:2_[M+H]+ | 2679 | -0.218153 | 0.015569 | 0.027245 |
| SM 40:1_[M+H]+ | 2697 | -0.212899 | 0.018246 | 0.030410 |
| SM 42:1_[M+H]+ | 2708 | -0.209689 | 0.020074 | 0.031936 |
| PC O-38:5_[M+H]+ | 2727 | -0.204144 | 0.023609 | 0.035926 |
| PC O-36:3_[M+H]+ | 2740 | -0.200350 | 0.026328 | 0.038394 |
| PC 40:6_[M+H]+ | 2754 | -0.196264 | 0.029554 | 0.041376 |

Statistical difference between ALS (n=77) and control (n=89) groups was assessed using the Mann-Whitney U test with Benjamini-Hochberg FDR correction for multiple comparisons. Uncorrected and corrected p-values are reported. Effect sizes are rank-biserial correlations (r, range -1 to +1), where negative values indicate higher abundance in ALS, 0 indicates complete overlap of distributions, and positive values indicate higher abundance in controls.

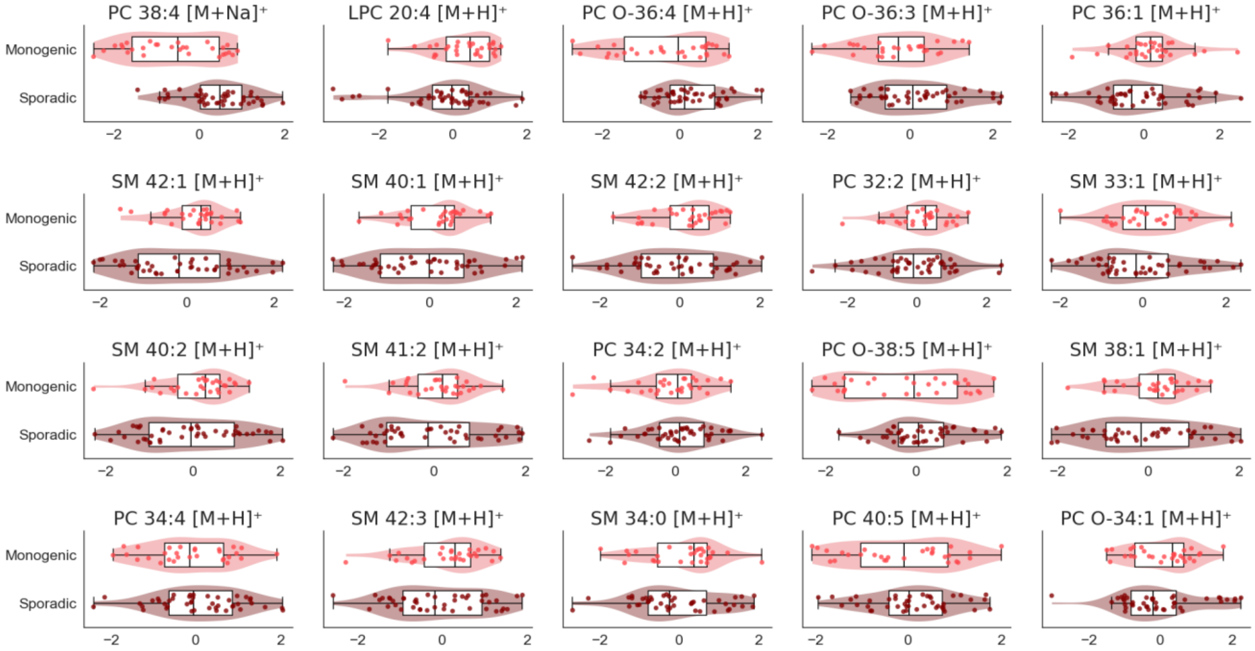


**Figure S1. Feature distributions for monogenic vs sporadic ALS.** Ranked by significance, although only PC 38:4 $[M+Na]^+$ differed significantly ($p$ = 0.005) between monogenic (n=29) and sporadic(n=44) ALS as determined by a Mann Whitney U test with Benjamini-Hochberg FDR correction for multiple comparisons.

**Table S6.** Statistical differences between monogenic and sporadic ALS.

| Feature | *U* statistic | Effect size | p-value | p-value (FDR) |
|---|---|---|---|---|
| PC 38:4_[M+Na]+ | 302 | -0.526646 | 0.000155 | 0.005441 |
| LPC 20:4_[M+H]+ | 831 | 0.302508 | 0.029999 | 0.524985 |
| PC O-36:4_[M+H]+ | 475 | -0.255486 | 0.066966 | 0.781266 |
| PC O-36:3_[M+H]+ | 502 | -0.213166 | 0.126630 | 0.786140 |
| PC 36:1_[M+H]+ | 770 | 0.206897 | 0.138225 | 0.786140 |
| SM 42:1_[M+H]+ | 753 | 0.180251 | 0.196777 | 0.786140 |
| SM 40:1_[M+H]+ | 746 | 0.169279 | 0.225560 | 0.786140 |
| SM 42:2_[M+H]+ | 735 | 0.152038 | 0.276653 | 0.786140 |
| PC 32:2_[M+H]+ | 733 | 0.148903 | 0.286730 | 0.786140 |
| SM 33:1_[M+H]+ | 731 | 0.145768 | 0.297052 | 0.786140 |
| SM 40:2_[M+H]+ | 728 | 0.141066 | 0.312995 | 0.786140 |
| SM 41:2_[M+H]+ | 726 | 0.137931 | 0.323932 | 0.786140 |
| PC 34:2_[M+H]+ | 550 | -0.137931 | 0.323932 | 0.786140 |
| PC O-38:5_[M+H]+ | 554 | -0.131661 | 0.346542 | 0.786140 |
| SM 38:1_[M+H]+ | 721 | 0.130094 | 0.352348 | 0.786140 |
| PC 34:4_[M+H]+ | 562 | -0.119122 | 0.394697 | 0.786140 |
| SM 42:3_[M+H]+ | 708 | 0.109718 | 0.433339 | 0.786140 |
| SM 34:0_[M+H]+ | 704 | 0.103448 | 0.460272 | 0.786140 |
| PC 40:5_[M+H]+ | 573 | -0.101881 | 0.467149 | 0.786140 |
| PC O-34:1_[M+H]+ | 702 | 0.100313 | 0.474083 | 0.786140 |

Top 20 lipids ranked by significance of difference between monogenic (n=29) and sporadic (n=44) ALS as determined by a Mann Whitney U test with Benjamini-Hochberg FDR correction for multiple comparisons. Effect sizes are rank-biserial correlations (r, range -1 to +1), where negative values indicate higher abundance in sporadic ALS, 0 indicates complete overlap of distributions, and positive values indicate higher abundance in monogenic ALS.

**Table S7.** Predictive model performance for classification of ALS vs control.

| Model | B/RS | F1 | Accuracy | Precision | Sensitivity | Specificity | AUCROC | AUPRC |
|---|---|---|---|---|---|---|---|---|
| LR | B | 0.79 (0.70, 0.87) | 0.81 (0.73, 0.88) | 0.81 (0.72, 0.90) | 0.77 (0.67, 0.86) | 0.84 (0.75, 0.93) | 0.90 (0.85, 0.94) | 0.90 (0.85, 0.95) |
| | RS | 0.83 (0.74, 0.93) | 0.85 (0.77, 0.93) | 0.85 (0.72, 0.97) | 0.83 (0.71, 0.95) | 0.85 (0.69, 1.00) | 0.92 (0.89, 0.96) | 0.93 (0.89, 0.97) |
| XGB | B | 0.73 (0.63, 0.83) | 0.75 (0.65, 0.85) | 0.73 (0.62, 0.83) | 0.74 (0.63, 0.86) | 0.75 (0.63, 0.87) | 0.88 (0.83, 0.92) | 0.87 (0.83, 0.91) |
| | RS | 0.83 (0.78, 0.88) | 0.85 (0.80, 0.89) | 0.85 (0.78, 0.91) | 0.83 (0.74, 0.91) | 0.87 (0.79, 0.94) | 0.94 (0.90, 0.98) | 0.93 (0.89, 0.98) |

**Abbreviations:** LR, Logistic Regression; XGB, XGBoost; B, Baseline; RS, Risk stratified.

Models trained on lipids only from the full cohort (n=166, $n_{ALS}$=77, $n_{HC}$=89). Risk stratified models removed unconfident predictions from evaluation. Metrics presented as mean (95% confidence interval).

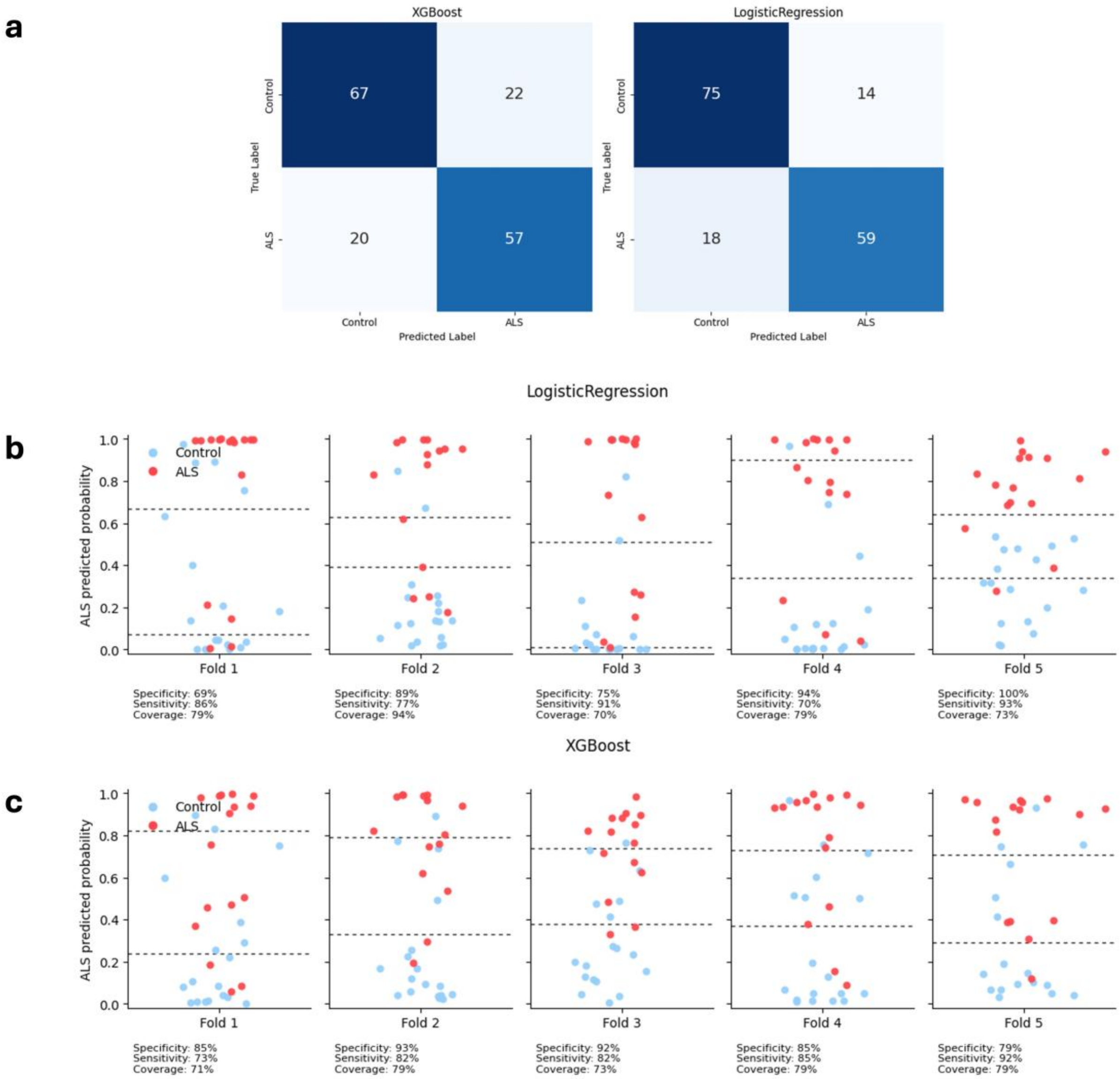


**Figure S2. Comparison of classification performance across modelling approaches.** Predictions are of models trained on lipid only data for all participants with ALS (n=77) or HC (n=89). (a) Confusion matrices demonstrating the number of correctly and incorrectly classified samples in XGBoost and Logistic Regression models. (b-c) Outer cross validation fold level predicted probabilities of ALS (y-axis) with optimised risk thresholds for each fold indicated by dashed lines. Dot colour indicates true diagnosis (blue=HC, red=ALS).

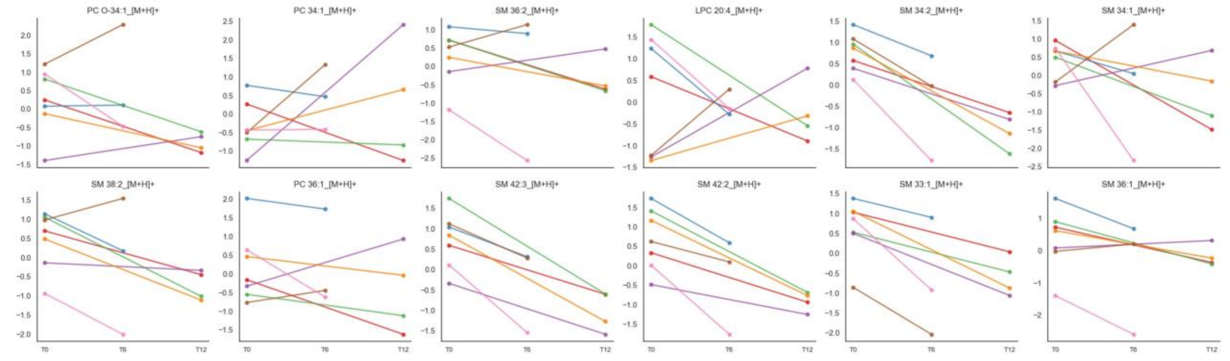


**Figure S3. Longitudinal trajectories of individual lipid features for each of the seven Tofersen-treated ALS patients**. The twelve features that differ most significantly between ALS and HC patients are presented. Panels display changes in selected lipid species contributing to the Logistic Regression model across timepoints for each patient, illustrating the feature-level dynamics underlying biomarker prediction risk trajectories. Distinct patterns are observed across individuals, with some patients showing coordinated shifts across multiple lipid features following treatment, whereas others exhibit relative stability or heterogeneous changes.

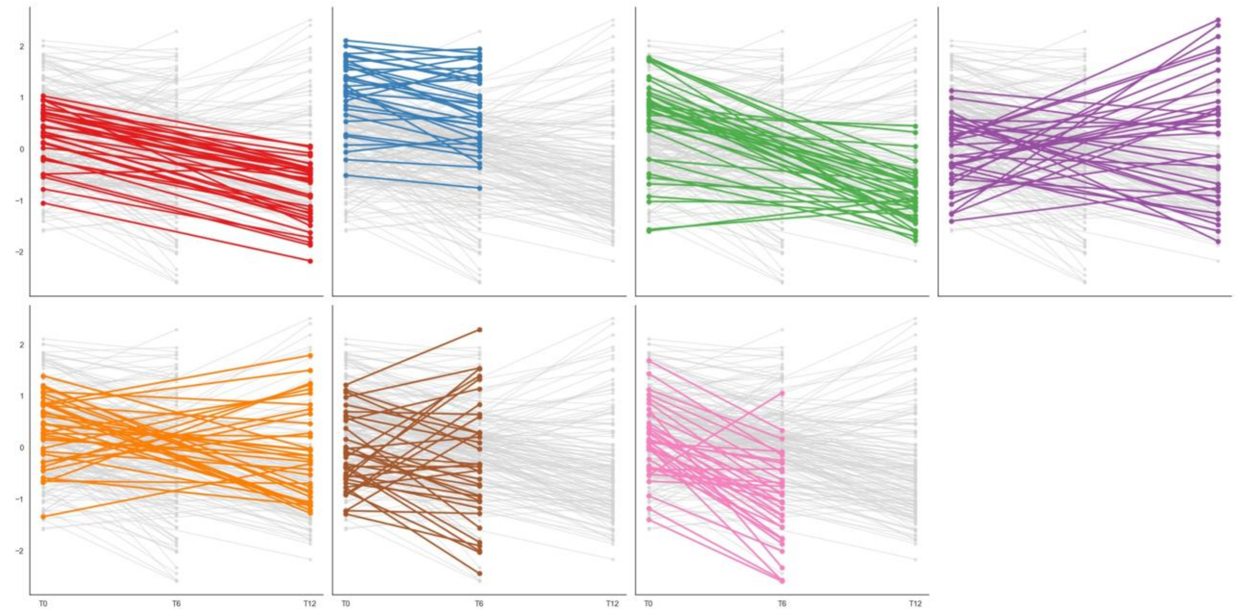

**Figure S4. Changes in all biomarkers for each Tofersen-treated ALS patient.** Heterogeneous patterns in biomarker change are observed in some patients, with more consistent directional shifts for others. These patient-specific trajectories demonstrate that changes in model-derived ALS probability arise from integrated, multi-feature alterations rather than single-analyte variation, supporting the biological basis of the biomarker and its sensitivity to inter-individual differences in disease state and treatment response.

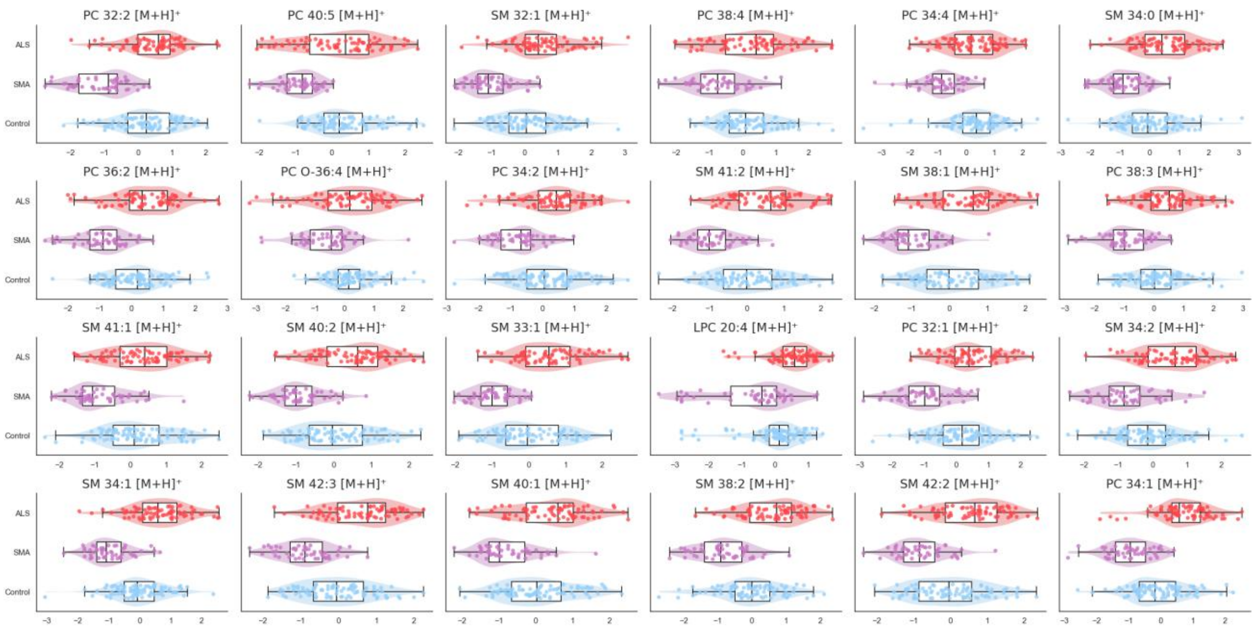


**Figure S5. Differential lipidomic distributions for individuals with SMA, ALS, and healthy controls.** Top 20 lipids that differ most significantly between SMA (n=50), ALS (n=77), and healthy controls (n=38) based on a Kruskal Wallis test with Benjamini-Hochberg correction.

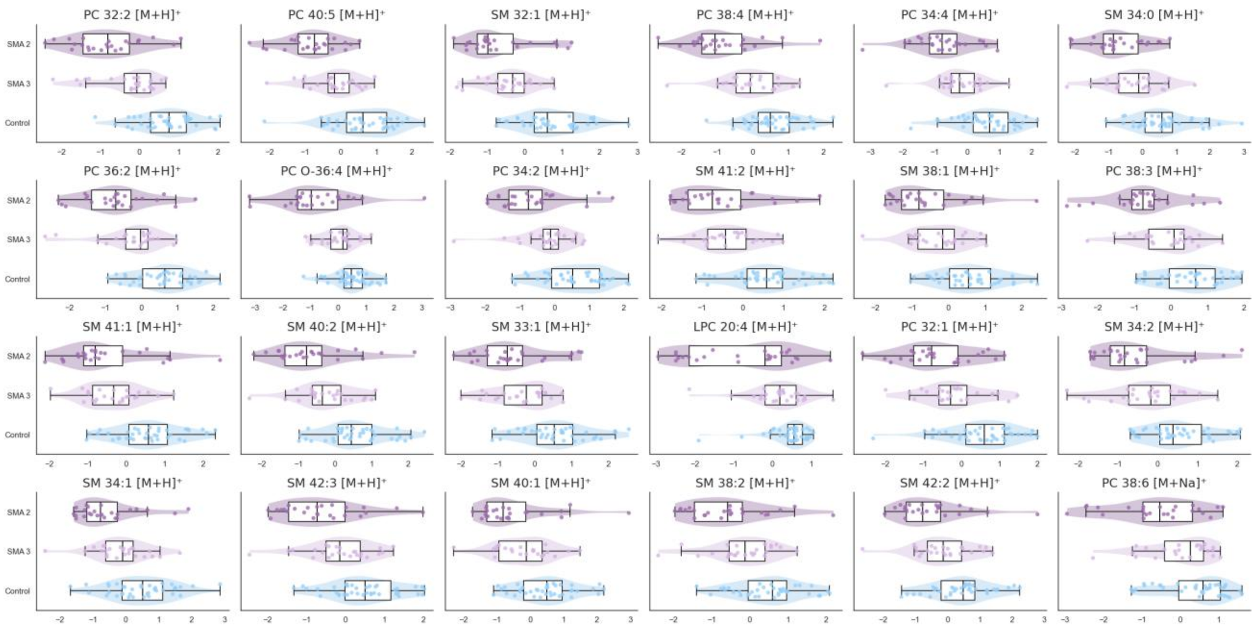


**Figure S6. Differential lipidomic distributions for individuals with SMA 2, SMA 3, and healthy controls.** Top 20 lipids that differ most significantly between SMA 2 (n=25), SMA 3 (n=25), and healthy controls (n=38) based on a Kruskal Wallis test with Benjamini-Hochberg correction.
